# Epidemiological and Clinical Characteristics of COVID-19 Patients with Cancers: A Systematic Review and Meta-Analysis of Global Data

**DOI:** 10.1101/2020.08.20.20177311

**Authors:** Xiangyi Kong, Yihang Qi, junjie Huang, Yang Zhao, Yongle Zhan, Xuzhen Qin, Zhihong Qi, Adejare (Jay) Atanda, Lei Zhang, Jing Wang, Yi Fang, Peng Jia, Asieh Golozar, Lin Zhang, Yu Jiang

## Abstract

**Summary:** Background Data on the prevalence of cancer in coronavirus disease 2019 (COVID-19)-infected patients and the severe illness incidence and mortality of COVID-19 patients with cancers remains unclear.

**Methods:** We systematically searched PubMed, Embase, Cochrane Library, and Web of Science, from database inception to July 15, 2020, for studies of patients with COVID-19 infection that had available comorbidity information on cancer. The primary endpoint was the pooled prevalence of cancer in COVID-19 patients and the secondary endpoint was the outcomes of COVID-19-infected cancer patients with incidence of severe illness and death rate. We calculated the pooled prevalence and corresponding 95% confidence intervals (95% CIs) using a random-effects model, and performed meta-regression analyses to explore heterogeneity. Subgroup analyses were conducted based on continent, country, age, sample size and study design.

**Findings:** A total of 107 eligible global studies were included in the systematic review. 90 studies with 94,845 COVID-19 patients in which 4,106 patients with cancer morbidity were included in the meta-analysis for prevalence of cancer morbidity among COVID-19 patients. 21 studies with 70,969 COVID-19 patients in which 3,351 patients with cancer morbidity who had severe illness or death during the studies. The overall prevalence of cancer among the COVID-19 patients was 0.07 (95% CI 0.05∽0.09). The cancer prevalence in COVID-19 patients of Europe (0.22, 95% CI 0.17∽0.28) was higher than that in Asia Pacific (0.04, 95% CI 0.03∽0.06) and North America (0.05, 95% CI 0.04∽0.06). The prevalence of COVID-19-infected cancer patients over 60 years old was 0.10 (95% CI 0.07∽0.14), higher than that of patients equal and less than 60 years old (0.05, 95% CI 0.03∽0.06). The pooled prevalence of severe illness among COVID-19 patients with cancers was 0.35 (95% CI 0.27∽0.43) and the pooled death rate of COVID-19 patients with cancers was 0.18 (95% CI 0.14∽0.18). The pooled incidence of severe illness of COVID-19 patients with cancers from Asia Pacific, Europe, and North America were 0.38(0.24, 0.52), 0.36(0.17, 0.55), and 0.26(0.20, 0.31), respectively; and the pooled death rate from Asia Pacific, Europe, and North America were 0.17(0.10, 0.24), 0.26(0.13, 0.39), and 0.19(0.13, 0.25), respectively.

**Interpretation:** To our knowledge, this study is the most comprehensive and up-to-date metaanalysis assessing the prevalence of cancer among COVID-19 patients, severe illness incidence and mortality rate. The prevalence of cancer varied significantly in geographical continents and age. The COVID-19 patients with cancer were at-risk for severe illness and a high death rate. The European COVID-19 patients had the highest cancer prevalence among the three continents examined and were also the most likely to progress to severe illness and death. Although the Asia Pacific COVID-19 patients had the lowest cancer prevalence, their severe illness rate was similar to that of European’s.

**Research in context:** 

**Evidence before this study:** Coronavirus disease (COVID-19) is an infectious disease caused by severe acute respiratory syndrome coronavirus 2 (SARS-CoV-2), a newly discovered coronavirus, which leads to respiratory illness and can be transmitted from person to person. As the infection has become widespread, concern for the influence of COVID-19 on patients with cancer has grown. Previous studies suggest that patients with a history of active malignancy might be at increased risk for COVID-19, developing COVID-19-related complications and having a poorer prognosis. Until now, however, few studies explored the following two questions: 1) what is the estimated prevalence of cancer patients with COVID-19 infection; and 2) do COVID-19-infected cancer-patients have distinct clinical courses and worse outcomes compared with COVID-19-infected patients without cancers. The latter is based on the former to further explore the characteristics of clinical outcomes of such patients. The clarification of these two questions will greatly help to understand the relationship between COVID-19 and cancer in terms of clinical epidemiology, and thus facilitate the formulation of targeted and relevant public health policies.

**Added value of this study:** To our knowledge, this systematic review and meta-analysis of 107 studies is the most comprehensive and up-to-date assessing the prevalence of cancer among COVID-19 patients, the incidence of severe illness and mortality rate of COVID-19 patients with cancers. We provided a relatively accurate overall cancer prevalence among the all COVID-19 patients (7%), stratified by geographical continent, country, age, study sample size, and study design type. We also presented the pooled severe illness and mortality rates stratified by continent. European COVID-19-infected cancer patients seemed the most likely to both develop cancer and progress to severe illness and death.

**Implications of all the available evidence:** Our findings have reinforced important considerations of clinical care and emphasized the urgent unmet needs for COVID-19 patients with cancers using the pooled prevalence, incidence of severe illness, and death rates as evidence. Also, after comparing the cancer prevalence, incidence of severe illness, and death rate of COVID-19 patients from different continents, European population may require stronger control measures than the Asia Pacific and North American populations. In the future, as more data will be available, it will be interesting to further investigate the differences of sociodemographic and climcopathological features between COVID-19-infected patients with cancer and without cancer.

## 1 Introduction

Globally, more than 18.5 million confirmed cases of coronavirus disease 2019 (COVID-19) has been reported (https://www.worldometers.info/coronavirus). Updated case counts are available on the World Health Organization (WHO) website. An interactive map highlighting confirmed cases throughout the world can be found at (https://coronavirus.jhu.edu/map.html). As infection has become widespread, concern for the influence of COVID-19 on patients with cancer has grown. Previous studies suggest that patients with a history of active malignancy might be at increased risk for COVID-19, developing COVID-19-related complications and having a poorer prognosis [1, 2], partly due to the following reasons [3]: *1)* a systemic immunosuppressive state caused by the malignancy and anticancer treatments, such as chemotherapy, surgery, or immunomodulatory drugs like PD-1/PD-L1 inhibitors [4–6]; *2)* older ages and major comorbidities including cancer, which increase the risk for COVID-19-related morbidity and mortality [7]; *3)* higher levels of contact with the healthcare system through provider visits for anticancer therapy, monitoring, and preventive and supportive care [8]. In February 2020, Liang et al. published a nationwide analysis of cancer patients with COVID-19 infection in China [9]. A total of 18 (1%) out of 1,590 COVID-19 patients had a cancer history, higher than the prevalence of cancer in the overall Chinese population (285·83per 100000 people [0·29%], based on 2015 cancer epidemiology statistics [10].

The clinical and prognostic characteristics of COVID-19-infected cancer patients have also been explored in recent studies. In March 2020, Zhang et al. carried out multivariate analyses on a total of 28 COVID-19-infected cancer patients from three hospitals in Wuhan, China, showing that cancer patients showed deteriorating conditions and poor outcomes from the COVID-19 infection [11]. Kuderer et al., on behalf of the COVID-19 and Cancer Consortium, found significant associations between 30-day all-cause mortality and the general factors of increasing age, male sex, former smoking, number of comorbidities, and receipt of azithromycin plus hydroxychloroquine [3]. These studies have however been restricted by small sample sizes, geographical area, and a lack of generalizability to the overall population of COVID-19 patients with cancer. In addition, limited clinical information, high heterogeneity of the course of the disease, as well as other critical issues render the epidemiological,clinical characteristics and treatment principles of COVID-19-infected cancer patients unclear. There is an urgent need to answer the following questions: *1)* what is the estimated prevalence of COVID-19-infected cancer patients as a function of geographic region, age, etc.?; *2)* Do COVID-19-infected cancer patients have distinct clinical courses and worse outcomes compared with COVID-19-infected patients without cancers; and *3)* Are there geographical differences in the severe illness and mortality rate among COVID-19-infected cancer patients. We answered these questions by conducting a systematic review and meta-analysis of observational studies on cancer patients in COVID-19 infection. Our findings will help in the management of COVID-19-infected cancer patients.

## 2 Methods

We followed the Preferred Reporting Items for Systematic Reviews and Meta-Analyses (PRISMA) guidelines to conduct this systematic review and meta-analysis [12]. Two reviewers (K.X. and Q.Y.) independently undertook the literature search, assessment for eligibility, data extraction and qualitative assessment. Inconsistencies were reviewed by a third reviewer (L.Z.) and resolved by consensus. The research protocol was registered and approved in PROSPERO (registration #CRD42020196014).

### 2.1 Data Sources and Searches

A comprehensive literature search of bibliographic databases was conducted to identify all relevant articles. To identify studies on COVID-19 in cancer patients, we searched PubMed, Embase, Cochrane Library and Web of Science from the inception of each database to June 31, 2020. Additionally, abstracts and presentations of all major conference proceedings were reviewed. Key/relevant MeSh terms and keywords included the following keywords: “2019-ncov”, “novel coronavirus”, “COVID-19”, “SARS-CoV-2”, “new coronavirus”, “coronavirus disease 2019”, “cancer”, “tumor”, “malignancy”, and “neoplasm”, etc. The exact literature search strategies are in **Table S1**. We also reviewed reference lists in a snow-ball technique to identify additional studies. Two investigators (K.X. and Q.Y.) independently screened titles and abstracts of identified articles. Major conflicts were resolved by a third researcher (Z.L.). Full text of identified studies were further reviewed by two independent reviewers. The search was again extended by review of references of articles included in the final selection.

### 2.2 Study Selection and Data Extraction

All studies containing the epidemiological and clinical information of COVID-19-infected cancer patients were identified with no geographic restrictions. Eligibility criteria included: *1)* studies reporting data on COVID-19 confirmed patients, survivors, and COVID-19 related death; *2)* studies containing available epidemiological or clinicopathological data of COVID-19-infected cancer patients; *3)* studies limited to humans. The exclusion criteria were as follows: *1)* letters, reviews, conference proceedings, commentaries, quality of life studies, cost-effectiveness analyses, and those in which the cancer data or COVID-19 data could not be ascertained. *2)* duplicate studies from same population or database (only the recent one to be included in the analysis). Two investigators (X.K. and Y.Q.) independently reviewed the list of retrieved articles to choose potentially relevant articles, and disagreements about studies were discussed and resolved with consensus. Both reviewers also independently extracted data from all studies and discrepancies were resolved in consensus with the third investigator L.Z. The following information was extracted from each publication: publication details, data collection period, sample size, sex, age, study design type, type of data used for characterization, clinical definition of COVID-19 as stated in the study, hospital where the cases were treated, patients’ basic vital signs, symptom and sign, race and ethnicity, smoking status, obesity status, number of comorbidities, time since cancer diagnosis to hospitalization, cancer histotype, tumor stage, treatments used for both cancer and COVID-19, laboratory results, severity staging, prognosis, etc.

### 2.3 Endpoint Setting and Stratification Strategy

Our primary outcome was prevalence of cancer among COVID-19 patients. The prevalence was defined as the number of COVID-19-infected cancer patients divided by the total number of COVID-19 patients in the study. The secondary outcomes included the incidence of severe illness and death rate among COVID-19-infected cancer patients. The stratification strategy for cancer prevalence in COVID-19 patients we adopted for the subgroup analysis was as follows:

1. by continent: patient populations in Europe, Asia Pacific, and North America were analyzed respectively.
2. by country: patient populations in China and other countries were analyzed respectively.
3. by age: We analyzed the cancer prevalence among COVID-19 patients stratified by mean age > 60 and mean age ≤ 60 years old.
4. by the sample size: we stratified the pooled prevalence of cancer in COVID-19 patients with sample size ≤100 and in larger studies with a sample size > 100.
5. by the study design type: we also stratified the pooled prevalence in cohort studies and non-cohort studies (case-series studies, case-control studies, and cross-sectional studies).

For incidence of severe illness and death rate of COVID-19 patients with cancer, the subgroup analysis stratified by continent was also performed to explore the difference in continents.

### 2.4 Data Synthesis and Analysis and Bias Risk Assessment

The statistical heterogeneity among studies was evaluated using Cochran’s Q test and I^2^ statistic, with I^2^ (% residual variation due to heterogeneity) values of 25%, 50%, and 75% representing low, moderate, and high heterogeneity, respectively [13]. If the I^2^ value was>50%, the random-effects model (FEM) would be applied. If the I^2^ value was < 50%, the fixed effects model (I’’EM) would be applied [14]. If substantial heterogeneity was detected, we performed meta-regression analyses to explore the proportion of between-study variance explained by participant characteristics and study characteristics. The pooled prevalence of cancer comorbidity in COVID-19 patient in different studies and corresponding 95% confidence intervals (CIs) were used to estimate the cancer prevalence among the COVID-19-infected population and the proportion of cancer patient with severe illness and death from COVID-19. We performed both Metan and Metaprop to pool the prevalence. Metaprop built further on the Metan procedure, which allowed computation of 95% CIs using the score statistic and the exact binomial method and incorporates the Freeman-Tukey double arcsine transformation of proportions[15]. The meta-analysis in Metaprop are shown in the main text and results of Metan are shown as a supplementary.

An estimation of publication bias was evaluated using Begg’s funnel plot, in which the SE of prevalence of each study was plotted against its prevalence and the corresponding Begg’s test was performed to test for small-study effects [16]. All reported P-values are 2-sided. A P-value less than 0.05 was considered to indicate statistical significance. All analyses were performed with Stata version 14.2 (StataCorp, College Station, TX, USA) [17, 18].

## 3 Results

### 3.1 Literature Search, Eligible Studies and Characteristics

**Figure 1** shows the study selection flowchart. After screening and eligibility assessment, we included a total of 107 studies (77 retrospective cohort studies, 22 case-series studies, 5 case-control studies, and 3 cross-sectional studies) in final quantitative and qualitative synthesis of evidence. Among the 107 studies, 90 studies reported the number of cancer comorbidity among COVID-19-infected patients published between December 2019 and June 2020 contained information on 94,845 COVID-19 patients in which 4,106 patients with cancer morbidity (**Table 1**). Among the 107 studies, 21 studies with 70,969 COVID-19 patients in which 3,351 patients with cancer morbidity who had severe illness or death during the studies provided detailed clinical outcome information of the COVID-19 patients with cancers, including illness severity and death to calculate the incidence of severe illness and death rate of COVID-19 patients with cancer (**Table 2**).

**Figure 1.**
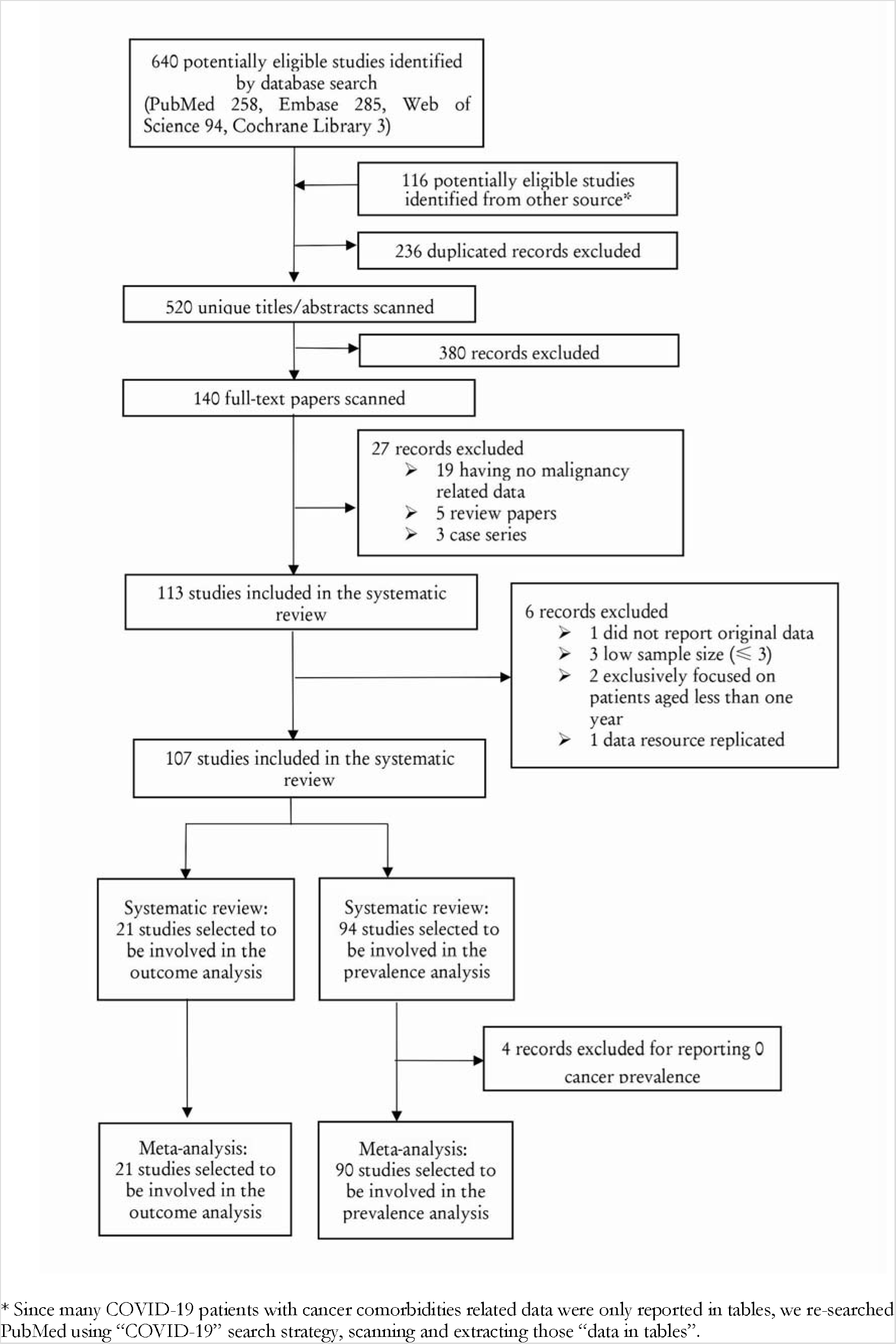
Study Exclusion and Inclusion Flowchart.

**Table 1.** The summary of characteristics of included studies of COVID-19 patients and cancer comorbidities [25] [26] [27] [28] [29][30] [31] [32] [33] [34] [35] [36] [37] [38] [39] [40] [41] [42] [43] [44] [45] [46] [47] [48] [49] [50] [51] [52] [53] [54] [55] [56] [57] [9] [58] [59] [60] [61] [62] [63] [64] [65] [66] [67] [68] [69] [70] [71] [72] [73] [74] [75] [75] [76] [77] [78] [79] [80] [81] [82] [83] [84] [85] [86] [87] [88] [89] [90] [91] [92] [93] [94] [95] [96] [97] [98] [99] [100] [101] [102] [103] [104] [105] [106] [107] [108] [109] [110] [111] [112] [113] [114] [115] [116] [117] [118].

| First author | Country | Continent | Study Design | Data Collection | Confirmation of COVID-19 | COVID-19 Patients | Age (years) | Male Percentage | Cancer |
| --- | --- | --- | --- | --- | --- | --- | --- | --- | --- |
| Beyrouti R[25] | UK | Europe | Case Series | Medical Records | RT-PCR | 6 | 61.0~85.0 | 0.83 | 2 |
| Cai Q[26] | China | Asia Pacific | Retrospective Cohort | EHR | RT-PCR | 298 | 47.5 ± 3.0~61.0 | 0.49 | 4 |
| Chen J[27] | China | Asia Pacific | Retrospective Cohort | EHR | Laboratory confirmed | 249 | 51.0 (36.0~64.0) | 0.51 | 1 |
| Chen L[28] | China | Asia Pacific | Retrospective Cohort | EHR | RT-PCR | 29 | 56.0 (26.0~79.0) | 0.72 | 1 |
| Chen N[29] | China | Asia Pacific | Retrospective Cohort | Medical Records | RT-PCR | 99 | 55.5 ± 13.1 | 0.68 | 1 |
| Chen T[30] | China | Asia Pacific | Case Series | EHR | RT-PCR | 274 | 62.0 (44.0~70.0) | 0.62 | 7 |
| Cheng Y[31] | China | Asia Pacific | Retrospective Cohort | Medical Records | RT-PCR | 701 | 63.0 (50.0~71.0) | 0.52 | 32 |
| Conversano A[32] | Italy | Europe | Retrospective Cohort | Medical Records | RT-PCR | 191 | 63.4 ± 14.9 | 0.69 | 29 |
| Dai M[33] | China | Asia Pacific | Retrospective Cohort | EHR | WHO Guideline | 641 | 64.0 ± 14.0 | 0.55 | 105 |
| Deng G[34] | China | Asia Pacific | Retrospective Cohort | EHR | RT-PCR | 44672 | NA | 0.51 | 107 |
| Du RH[35] | China | Asia Pacific | Retrospective Cohort | EHR | RT-PCR | 109 | 70.7 ± 10.9 | 0.68 | 8 |
| Du Y[36] | China | Asia Pacific | Retrospective Cohort | EHR | PCR | 85 | 65.8 ± 14.2 | 0.73 | 6 |
| Duanmu Y[37] | USA | North America | Cross-Sectional Study | EHR | PCR | 100 | 45.0 (32.0~65.0) | 0.56 | 3 |
| Feng Y[38] | China | Asia Pacific | Retrospective Cohort | Medical Records | RT-PCR | 476 | 53.0 (40.0~64.0) | 0.57 | 57 |
| Fernández-Ruiz M[39] | Spain | Europe | Case Series | Medical Records | RT-PCR | 18 | 71.0 ± 12.8 | 0.78 | 4 |
| Grasselli G[40] | Italy | Europe | Case Series | EHR | RT-PCR | 1591 | 63.0 (56.0~70.0) | 0.82 | 81 |
| Grillet F[41] | France | Europe | Retrospective Cohort | Medical Records | RT-PCR | 100 | 66.0 ± 13.0 | 0.70 | 20 |
| Guan WJ[42] | China | Asia Pacific | Retrospective Cohort | EHR | RT-PCR | 1590 | 49.0 ± 16.0 | 0.57 | 130 |
| Guan WJ[43] | China | Asia Pacific | Retrospective Cohort | EHR | RT-PCR | 1099 | 47.0 (35.0~58.0) | 0.58 | 10 |
| Guo W[44] | China | Asia Pacific | Retrospective Cohort | EHR | RT-PCR | 174 | 59.0 (49.0~67.0) | 0.44 | 21 |
| He Y[45] | China | Asia Pacific | Retrospective Cohort | Medical Records | Not specified | 65 | 51.0 (27.0~68.0) | 0.48 | 1 |
| Hou W[46] | China | Asia Pacific | Retrospective Cohort | EHR | PCR | 101 | 50.9 ± 20.1 | 0.44 | 5 |
| Hu L[47] | China | Asia Pacific | Retrospective Cohort | Medical Records | RT-PCR | 323 | 61.0 (23.0~91.0) | 0.51 | 5 |
| Huang C[48] | China | Asia Pacific | Retrospective Cohort | EHR | RT-PCR | 41 | 49.0 (41.0~58.0) | 0.73 | 1 |
| Ihle-Hansen H[49] | Norway | Europe | Retrospective Cohort | Medical Records | RT-PCR | 43 | 67.8 | 0.67 | 5 |
| Inciardi R M[50] | Italy | Europe | Retrospective Cohort | Medical Records | RT-PCR | 99 | 68.0 ± 12.0 | 0.85 | 33 |
| Ji M[51] | China | Asia Pacific | Retrospective Cohort | EHR | RT-PCR | 101 | 51.0 (37.0~61.0) | 0.48 | 12 |
| Jin X[52] | China | Asia Pacific | Retrospective Cohort | Medical Records | RT-PCR | 651 | 46.1 ± 14.2 | NA | 0 |
| KCDC[53] | Korea | Asia Pacific | Cross-Sectional Study | Government reports | NA | 54 | 75.5 (66.0~80.0) | 0.61 | 13 |
| Kalligeros M[54] | USA | North America | Retrospective Cohort | EHR | RT-PCR | 103 | 60.0 (52.0~70.0) | 0.61 | 9 |
| Li J[55] | China | Asia Pacific | Case Series | Medical Records | RT-PCR | 1178 | 55.5 (38.0~67.0) | 0.46 | 32 |
| Li K[56] | China | Asia Pacific | Retrospective Cohort | Medical Records | RT-PCR | 78 | 44.6 | 0.49 | 9 |
| Li X[57] | China | Asia Pacific | Retrospective Cohort | EHR | WHO Guideline | 548 | 60.0 (48.0~69.0) | 0.51 | 24 |
| Liang W[9] | China | Asia Pacific | Retrospective Cohort | EHR | Laboratory confirmed | 1590 | NA | NA | 18 |
| Lin L[58] | China | Asia Pacific | Case-Control Study | Medical Records | RT-PCR | 95 | 45.3 | 0.47 | 5 |
| Liu K[59] | China | Asia Pacific | Retrospective Cohort | EHR | RT-PCR | 137 | 57.0 (20.0~83.0) | 0.45 | 2 |
| Liu W[60] | China | Asia Pacific | Retrospective Cohort | Medical Records | RT-PCR | 78 | 38.0 (33.0~57.0) | 0.64 | 4 |
| Liu Y[61] | China | Asia Pacific | Case Series | Medical Records | Laboratory confirmed | 12 | NA | 0.66 | 0 |
| Lovell N[62] | UK | Europe | Case Series | Medical Records | Not specified | 101 | 82.0 (72.0~89.0) | NA | 25 |
| Luong-Nguyen M[63] | France | Europe | Retrospective Cohort | Medical Records | Not specified | 15 | 62.0 (35.0~68.0) | 0.60 | 8 |
| Malard F[64] | France | Europe | Retrospective Cohort | Medical Records | PCR | 25 | 72.0 | 0.68 | 20 |
| Mancia G[65] | Italy | Europe | Case-Control Study | Medical Records | RT-PCR | 6272 | 68.0 ± 13.0 | 0.63 | 1091 |
| Mao L[66] | China | Asia Pacific | Case Series | EHR | RT-PCR | 214 | 52.7 ± 15.5 | 0.41 | 13 |
| Meng Y[67] | China | Asia Pacific | Retrospective Cohort | Medical Records | Not specified | 168 | 67.0 ± 15.0 | 0.52 | 1 |
| Million M[68] | France | Europe | Retrospective Cohort | EHR | RT-PCR | 1061 | 43.6 | 0.46 | 161 |
| Miyashita H[69] | USA | North America | Retrospective Cohort | EHR | RT-PCR | 5688 | NA | NA | 334 |
| Molina JM[70] | France | Europe | Retrospective Cohort | Medical Records | PCR | 11 | 58.7 | 0.64 | 5 |
| Montopoli M[71] | Italy | Europe | Case-Control Study | EHR | WHO Guideline | 9280 | NA | 0.49 | 786 |
| Myers LC[72] | USA | North America | Retrospective Cohort | Medical Records | PCR | 377 | 61.0 (50.0~73.0) | 0.56 | 18 |
| Nair V[73] | USA | North America | Case Series | EHR | RT-PCR | 10 | 57.0 (47.0~67.0) | 0.60 | 0 |
| O'Reilly GM[74] | Australia | Asia Pacific | Retrospective Cohort | EHR | RT-PCR | 240 | 60.0 ± 21.0 | 0.55 | 1 |
| Pan L[75] | China | Asia Pacific | Cross-Sectional Study | EHR | RT-PCR | 204 | 52.9 ± 16.0 | 0.55 | 13 |
| Pereira MR[76] | USA | North America | Case Series | Medical Records | RT-PCR | 90 | 57.0 (46.0~68.0) | 0.59 | 3 |
| Qi X[77] | China | Asia Pacific | Retrospective Cohort | Medical Records | Not specified | 70 | 41.0 (27.5~50.0); | 0.72 | 2 |
| Qin C[78] | China | Asia Pacific | Retrospective Cohort | EHR | WHO Guideline | 452 | 58.0 (22.0~95.0) | 0.52 | 54 |
| Safiya Richardson[79] | USA | North America | Case Series | EHR | PCR | 5700 | 63.0 (52.0~75.0) | 0.60 | 320 |
| Shi H[80] | China | Asia Pacific | Retrospective Cohort | EHR | RT-PCR | 81 | 49.5 ± 11.0 | 0.52 | 4 |
| Shi S[81] | China | Asia Pacific | Retrospective Cohort | Medical Records | WHO Guideline | 416 | 64.0 (21.0~95.0) | 0.49 | 9 |
| Shi Y[82] | China | Asia Pacific | Retrospective Cohort | EHR | WHO Guideline | 487 | 46.0 ± 19.0 | 0.53 | 5 |
| Stroppa EM[83] | Italy | Europe | Retrospective Cohort | EHR | RT-PCR | 56 | 71.64 (50~84) | 0.8 | 25 |
| Sun B[84] | China | Asia Pacific | Retrospective Cohort | Medical Records | Not specified | 38 | 58.0 (49.0~69.5) | .91 | 4 |
| Sun H[85] | USA | North America | Case Series | EHR | PCR | 30 | 84.5 (71.0~97.0) | 0.47 | 7 |
| Sun Y[86] | Singapore | Singapore | Case-Control Study | EHR | PCR | 788 | 34.0 | 0.49 | 94 |
| Wan S[87] | China | Asia Pacific | Case Series | EHR | RT-PCR | 135 | 47.0 (36.0~5.0) | 0.53 | 4 |
| Wang B[88] | China | Asia Pacific | Case Series | Medical Records | RT-PCR | 26 | 5.0~72.0 | 0.42 | 1 |
| Wang D[89] | China | Asia Pacific | Case Series | EHR | RT-PCR | 138 | 56.0 (42.0-68.0) | 0.54 | 10 |
| Wang K[90] | China | Asia Pacific | Case Series | Medical Records | RT-PCR | 144 | 53.0 | 0.51 | 1 |
| Wang L[91] | China | Asia Pacific | Case Series | EHR | PCR | 26 | 42.0 (33.5~53.3) | 0.42 | 1 |
| Wang L[92] | China | Asia Pacific | Retrospective Cohort | Medical Records | RT-PCR | 116 | 54.0 (38.0~69.0) | 0.58 | 12 |
| Wang L[93] | China | Asia Pacific | Retrospective Cohort | EHR | Laboratory confirmed | 339 | 71.0 ± 8.0 | NA | 15 |
| Wang Y[94] | China | Asia Pacific | Retrospective Cohort | Medical Records | WHO Guideline | 46 | NA | 0.57 | 2 |
| Wang Z[95] | China | Asia Pacific | Case Series | EHR | RT-PCR | 69 | 42.0 (35.0~62.0) | 0.46 | 4 |
| Wang K[96] | China | Asia Pacific | Retrospective Cohort | EHR | Not specified | 114 | 53.0 (23.0~78.0) | 0.51 | 1 |
| Wu C[97] | China | Asia Pacific | Retrospective Cohort | EHR | RT-PCR | 201 | 51.0 (43.0~60.0) | 0.64 | 1 |
| Wu J[98] | China | Asia Pacific | Retrospective Cohort | EHR | WHO Guideline | 280 | 43.1 ± 19.0 | 0.54 | 33 |
| Xie J[99] | China | Asia Pacific | Retrospective Cohort | Medical Records | RT-PCR | 140 | 60.0 (47.0~68.0) | 0.51 | 5 |
| Xu X[100] | China | Asia Pacific | Case Series | EHR | RT-PCR | 90 | 50.0 (18.0~86.0) | 0.43 | 2 |
| Yang F[101] | China | Asia Pacific | Retrospective Cohort | Medical Records | RT-PCR | 92 | 69.8 (30.0~97.0) | 0.58 | 4 |
| Yang F[102] | China | Asia Pacific | Retrospective Cohort | EHR | RT-PCR | 1575 | 63 (34~98) | 0.54 | 52 |
| Yang X[103] | China | Asia Pacific | Retrospective Cohort | EHR | Laboratory confirmed | 52 | 59.7 (13.3) | 0.67 | 2 |
| Yao Q[104] | China | Asia Pacific | Retrospective Cohort | Medical Records | Not specified | 108 | 52.0 (37.0~58.0) | 0.40 | 2 |
| Yasukawa K[105] | USA | North America | Retrospective Cohort | Medical Records | Not specified | 50 | 62.0 (22.0~78.0) | 0.58 | 4 |
| Yin L[106] | China | Asia Pacific | Case-Control Study | EHR | RT-PCR | 45 | 52.4 | 0.56 | 3 |
| Yu Q[107] | China | Asia Pacific | Retrospective Cohort | Medical Records | Laboratory confirmed | 625 | 46.9 ± 15.4 | 0.53 | 6 |
| Yuan M[108] | China | Asia Pacific | Retrospective Cohort | Medical Records | RT-PCR | 27 | 60.0 (47.0~69.0) | 0.44 | 1 |
| Zhang J[109] | China | Asia Pacific | Retrospective Cohort | Medical Records | RT-PCR | 290 | NA | 0.53 | 35 |
| Zhang J[110] | China | Asia Pacific | Retrospective Cohort | Medical Records | Laboratory confirmed | 111 | 38.0 (32.0~57.0) | 0.41 | 8 |
| Zhang L[111] | China | Asia Pacific | Retrospective Cohort | EHR | Laboratory confirmed | 343 | 62.0 (48.0~69.0) | 0.50 | 41 |
| Zhang R[112] | China | Asia Pacific | Case Series | EHR | RT-PCR | 120 | 45.4 ± 15.6 | 0.36 | 7 |
| Zhang X[113] | China | Asia Pacific | Retrospective Cohort | EHR | WHO Guideline | 645 | 46.7 ± 13.8 | 51.5% | 0 |
| Zhou F[114] | China | Asia Pacific | Retrospective Cohort | EHR | RT-PCR | 191 | 56.0 (18.0~87.0) | 0.62 | 2 |
| Zhou Z[115] | China | Asia Pacific | Retrospective Cohort | EHR | WHO Guideline | 254 | 50.6 (5.0~87.0) | 0.45 | 30 |
| Zhu W[116] | China | Asia Pacific | Retrospective Cohort | EHR | PCR | 32 | 46.0 (35.0~52.0) | 0.47 | 2 |
| Ziehr DR[117] | USA | North America | Retrospective Cohort | EHR | Not specified | 66 | 58.0 (23.0~87.0) | 0.65 | 5 |

Note:
COVID-19 Definition from WHO Guideline [118]
Age (year): Mean± SD or Median (range)

**Table 2.** The summary of characteristics of included studies of CO VID-19 with cancer and clinical outcome [33] [42] [119] [9] [120] [121] [122] [123] [69] [71] [8] [124] [83] [125] [126] [102] [127] [128] [129] [130].

COVID-19 Definition from WHO Guideline [118] Age (year): Mean± SD or Median (range)
| Table 2 The summary of characteristics of included studies of COVID-19 with cancer and clinical outcome |  |  |  |  |  |  |  |  |  |  |  |  |
| --- | --- | --- | --- | --- | --- | --- | --- | --- | --- | --- | --- | --- |
| First author | Country | Study Design | Data Collection | COVID-19 Confirmation | COVID-19 Patients | Age (year) | Male Percentage | Cancer Patients | Severe illness with cancer | Severe illness without cancer | Death with cancer | Death without cancer |
| Dai M[33] | China | Retrospective Cohort | EHR | WHO guideline | 641 | 64 | 0.55 | 105 | 35 | 75 | 12 | 27 |
| Guan WJ[42] | China | Retrospective Cohort | EHR | RT-PCR | 1590 | 49 | 0.57 | 130 | 9 | 245 | 3 | 47 |
| He W[119] | China | Retrospective Cohort | EHR | CT scan | 13 | 35 | 0.54 | 13 | 4 | NA | 8 | NA |
| Liang W[9] | China | Retrospective Cohort | EHR | Laboratory confirmed | 1590 | 63.1 | NA | 18 | 7 | 124 | NA | NA |
| Luo J[120] | USA | Retrospective Cohort | EHR | RT-PCR | 69 | 69 (31~91) | 0.48 | 69 | 24 | NA | 16 | NA |
| Ma J[121] | China | Retrospective Cohort | EHR | RT-PCR | 37 | 62 | 0.541 | 37 | 20 | NA | 5 | NA |
| Martin-MoroF[122] | Spain | Retrospective Cohort | EHR | RT-PCR | 34 | 72.5 (35~94) | 0.56 | 34 | NA | NA | 11 | NA |
| Mehta V[123] | USA | Retrospective Cohort | EHR | RT-PCR | 218 | 69(10~92) | NA | 218 | 45 | NA | 61 | NA |
| Miyashita H[69] | USA | Retrospective Cohort | EHR | RT-PCR | 5688 | NA | NA | 334 | NA | NA | 37 | 518 |
| Montopoli M[71] | Italy | Case-Control Study | EHR | WHO guideline | 4532 | NA | 0.49 | 430 | 121 | NA | 75 | 312 |
| Nicole M Kuderer[8] | USA, Canada and Spain | Retrospective Cohort | EHR | Laboratory confirmed | 928 | 66 (57~76) | 0.5 | 928 | 242 | NA | 121 | NA |
| Rafi Kabarriti NPB[124] | USA | Retrospective Cohort | EHR | RT-PCR | 107 | 70 (30~95) | 0.5 | 107 | NA | NA | 24 | NA |
| Stroppa EM[83] | Italy | Retrospective Cohort | EHR | RT-PCR | 56 | 71 (50~84) | 0.8 | 25 | 12 | NA | 9 | NA |
| TNAde Rojas[125] | Spain | Case Series | EHR | PCR | 15 | 11(1~19) | 0.93 | 15 | NA | NA | NA | NA |
| Wei X[126] | China | Retrospective Cohort | EHR | RT-PCR | 252 | 64.8 ± 13.3 | 0.52 | 252 | 121 | NA | NA | NA |
| Yang F[102] | China | Retrospective Cohort | EHR | RT-PCR | 1575 | 63 (34~98) | 0.54 | 52 | 19 | NA | 11 | NA |
| Yang KY[127] | China | Retrospective Cohort | EHR | RT-PCR | 8161 | 63 (14~96) | 0.47 | 205 | 52 | NA | 40 | NA |
| Yu J[128] | China | Case Series | EHR | WHO guideline | 12 | 66 | 0.83 | 12 | 3 | NA | 3 | NA |
| Zhang L[129] | China | Retrospective Cohort | EHR | RT-PCR | 28 | 65 | 0.61 | 28 | 15 | NA | 8 | NA |
| Tian J[130] | China | Retrospective Cohort | EHR | WHO guideline | 751 | 64 | 0.5 | 232 | 148 | 166 | 46 | 56 |

Note: COVID-19 Definition from WHO Guideline [118] Age (year): Mean±SD or Median (range)

### 3.2 Prevalence of Cancers among COVID-19 Patients

The overall cancer prevalence among the entire COVID-19 patient cohort was 0.07 (95% CI 0.05∼0.09, weights were from random-effects analysis model, **Figure 2**). For subgroup analysis, patient population was from Europe (14 studies in total, including 6 from Italy, 4 from France, 2 from the UK, 1 from Spain, and 1 from Norway), Asia Pacific (72 studies in total, including 70 from China and 1 from Korea, 1 study from Australia), and North America (all 8 studies were from the USA). There were 36 studies with patients over 60 years old; for the other 55 studies, patients’ were under 60 years old. Results of subgroup analysis showed that the prevalence of COVID-19 among cancer patients in Europe (prevalence rate 0.22, 95% CI 0.17∽0.28) was higher than that in Asia Pacific (0.04, 95% CI 0.03∽0.06) and North America (0.05, 95% CI 0.04∽0.06) (**Figure 3**). If stratified by the country, the prevalence of COVID-19-infected cancer was 0.04 (95%. CI 0.03∽0.06) in China, lower than that in other countries (0.13, 95% CI 0.11∽0.16) (**Figure 4**). The prevalence of COVID-19-infected cancer patients over 60 years old was 0.10 (95% CI 0.07∽0.14), higher than that of patients <=60 years old (0.05, 95% CI 0.03∽0.06) (**Figure 5**). The prevalence was slightly higher, at 0.10 (95% CI 0.07∽0.14) in studies with a sample size <=100, but in larger studies with a sample size > 100, the overall prevalence was lower at 0.05 (95% CI 0.04∽0.07) (**Figure 6**). As shown in **Figure 7**, the pooled prevalence upon cohort studies was also estimated to be 0.07 (95% CI 0.05∽0.09), equal to the prevalence based on the included 26 non-cohort studies (0.07, 95% CI 0.05∽0.09). The results obtained from the Metan method (**Figure S1A-F** in supplementary material) were similar as presented in Metaprop method.

**Figure 2.**
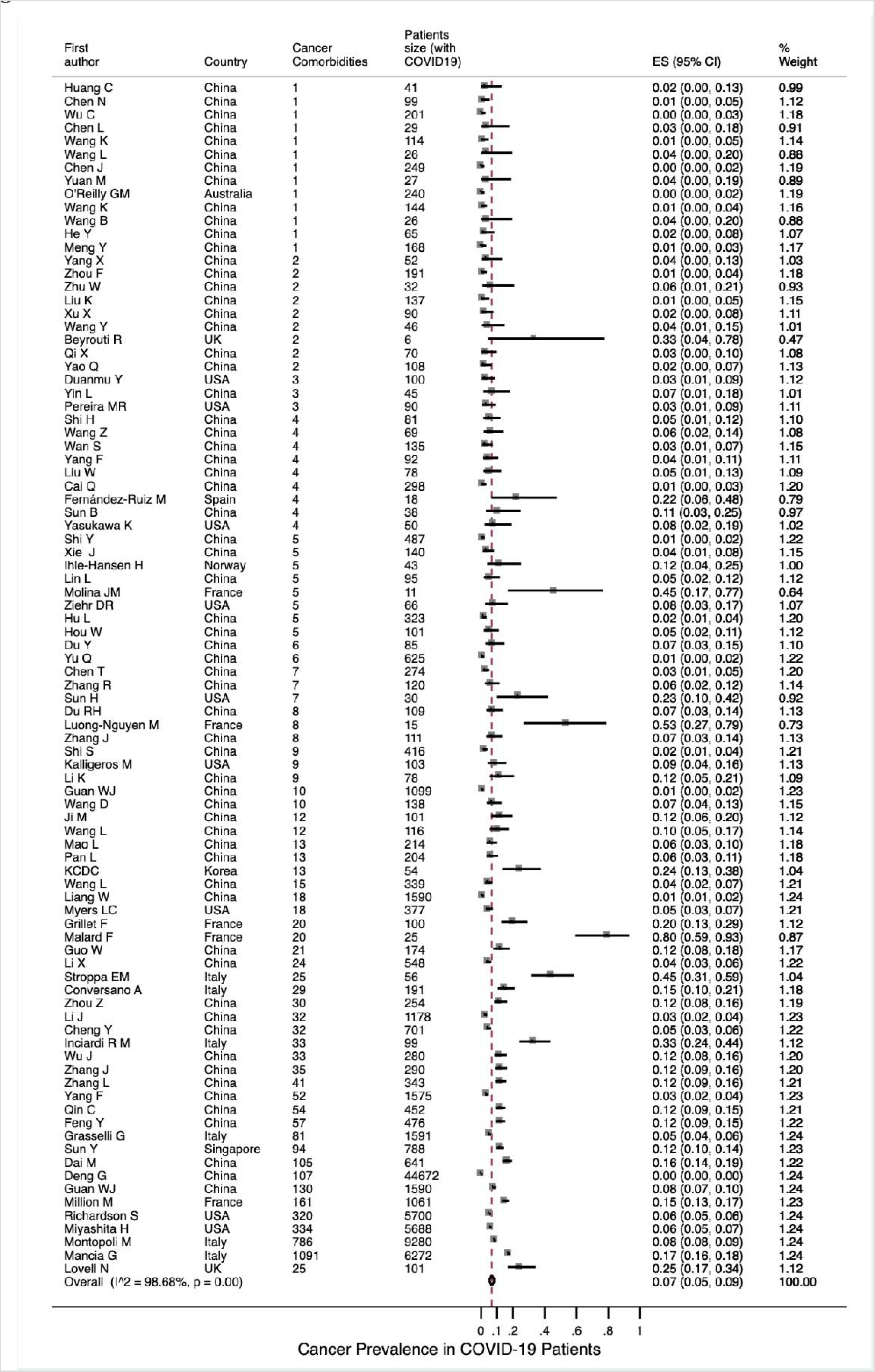
Overall Cancer Prevalence in COVID-19 Patients

**Figure 3.**
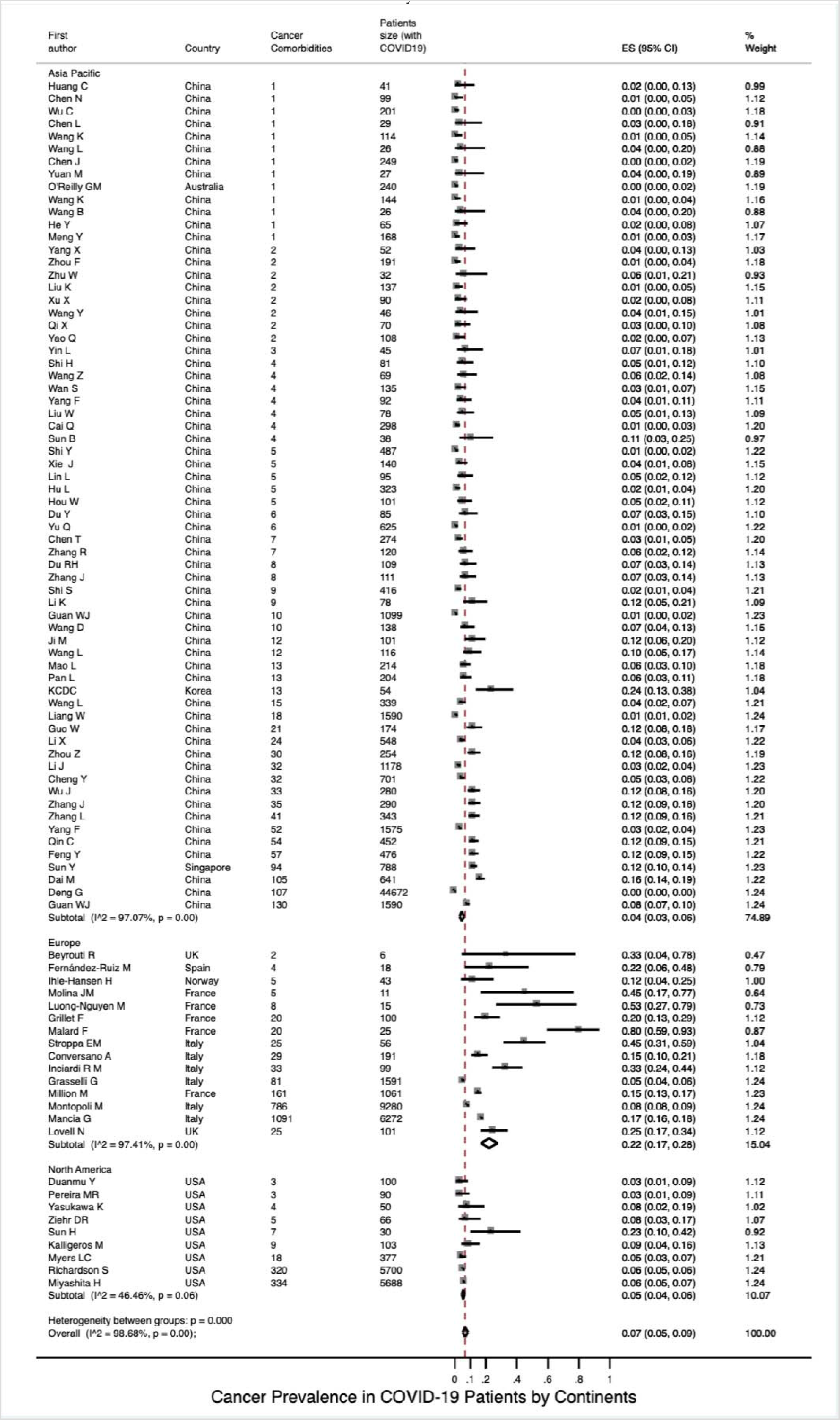
Cancer Prevalence in COVID-19 Patients by Continents

**Figure 4.**
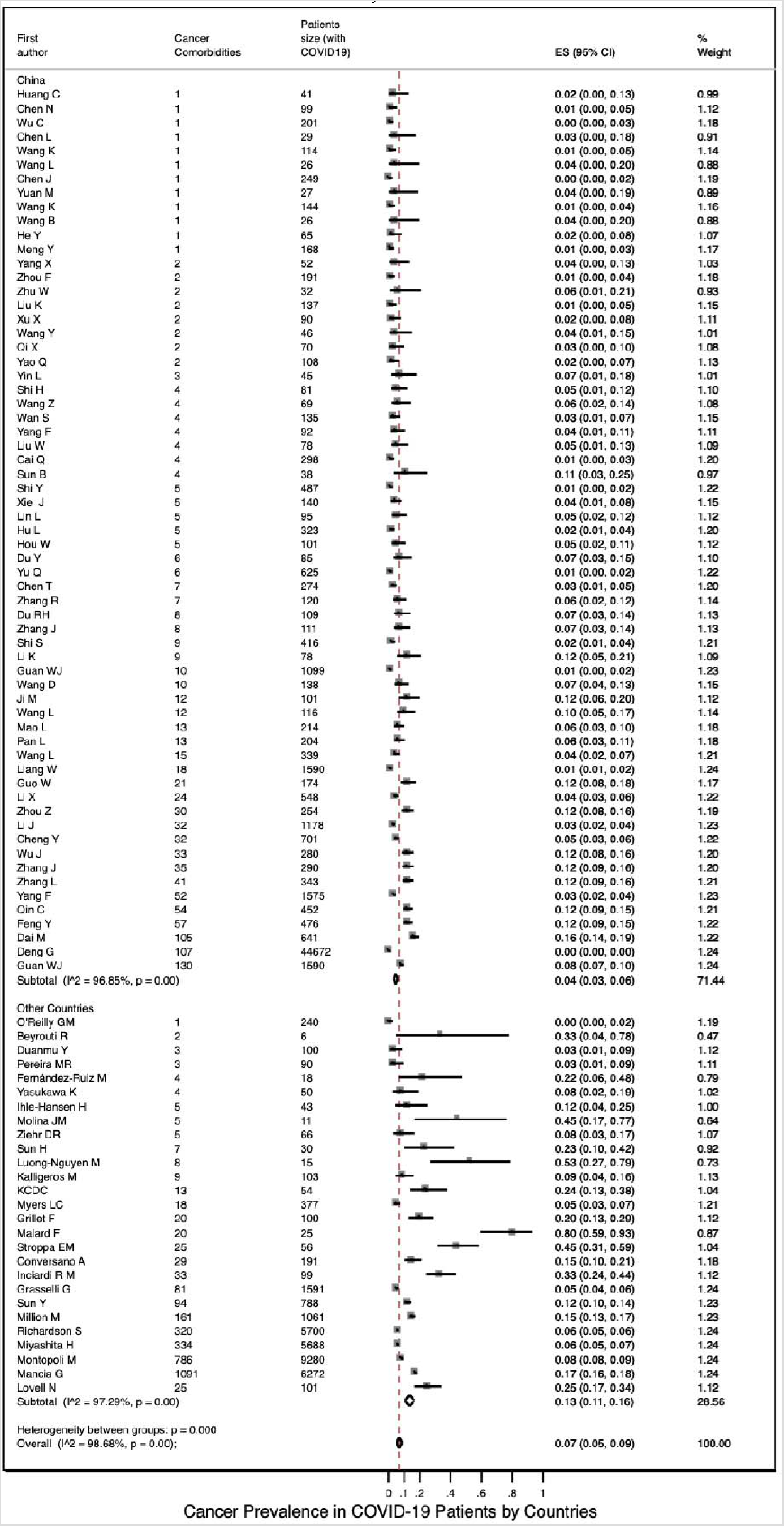
Cancer Prevalence in COVID-19 Patients by Countries

**Figure 5.**
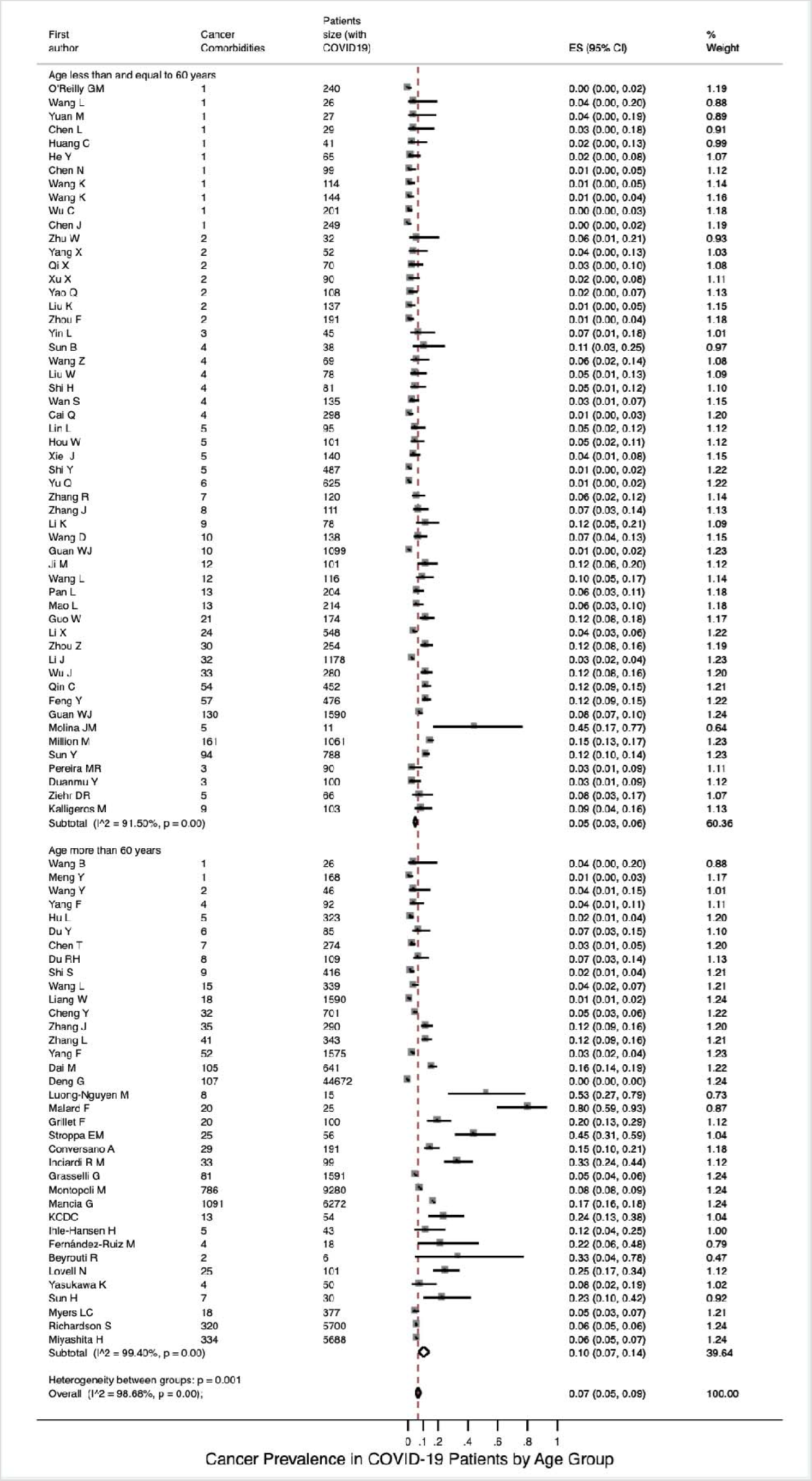
Cancer Prevalence in COVID-19 Patients by Age Group

**Figure 6.**
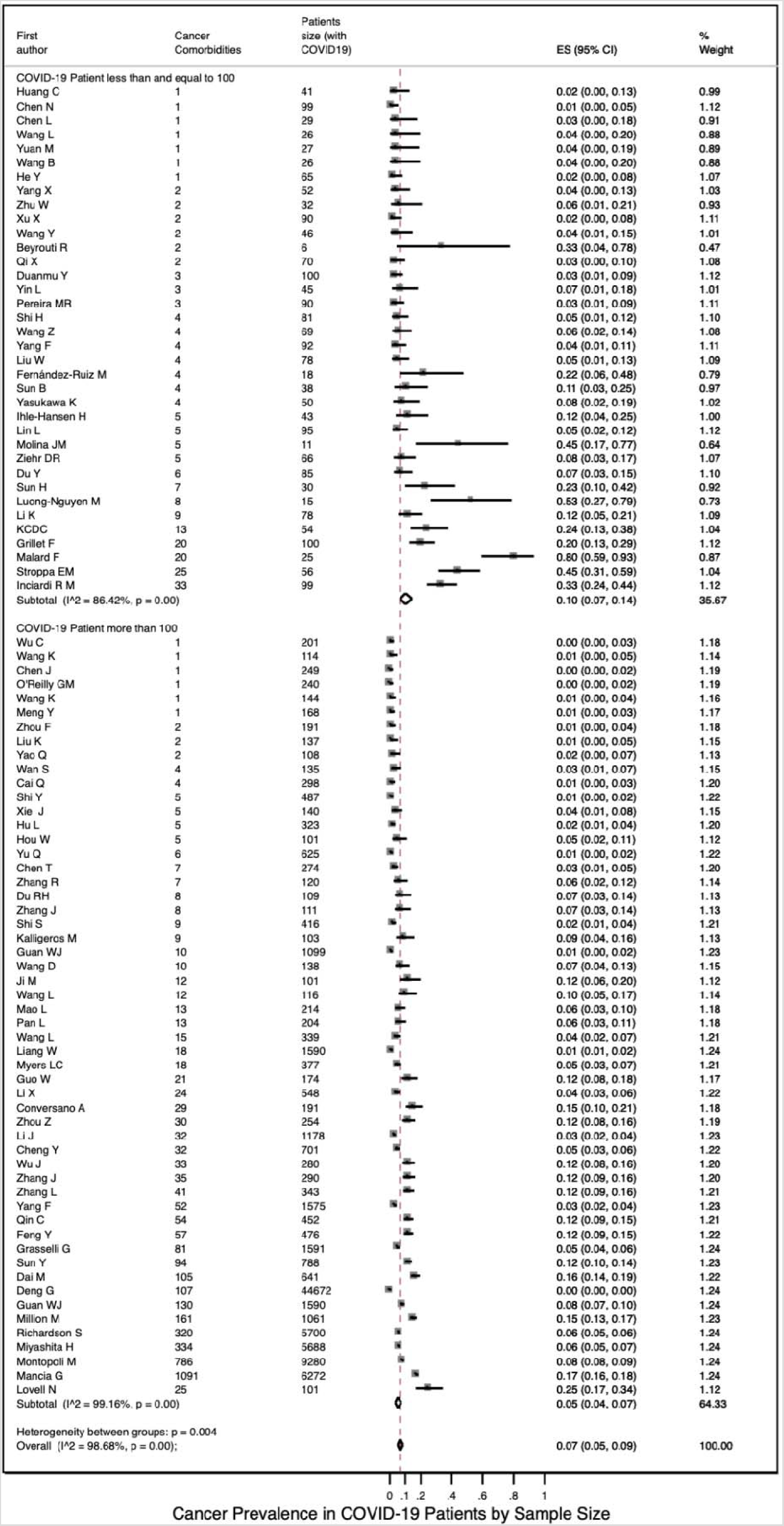
Cancer Prevalence in COVID-19 Patients by Sample Size

**Figure 7.**
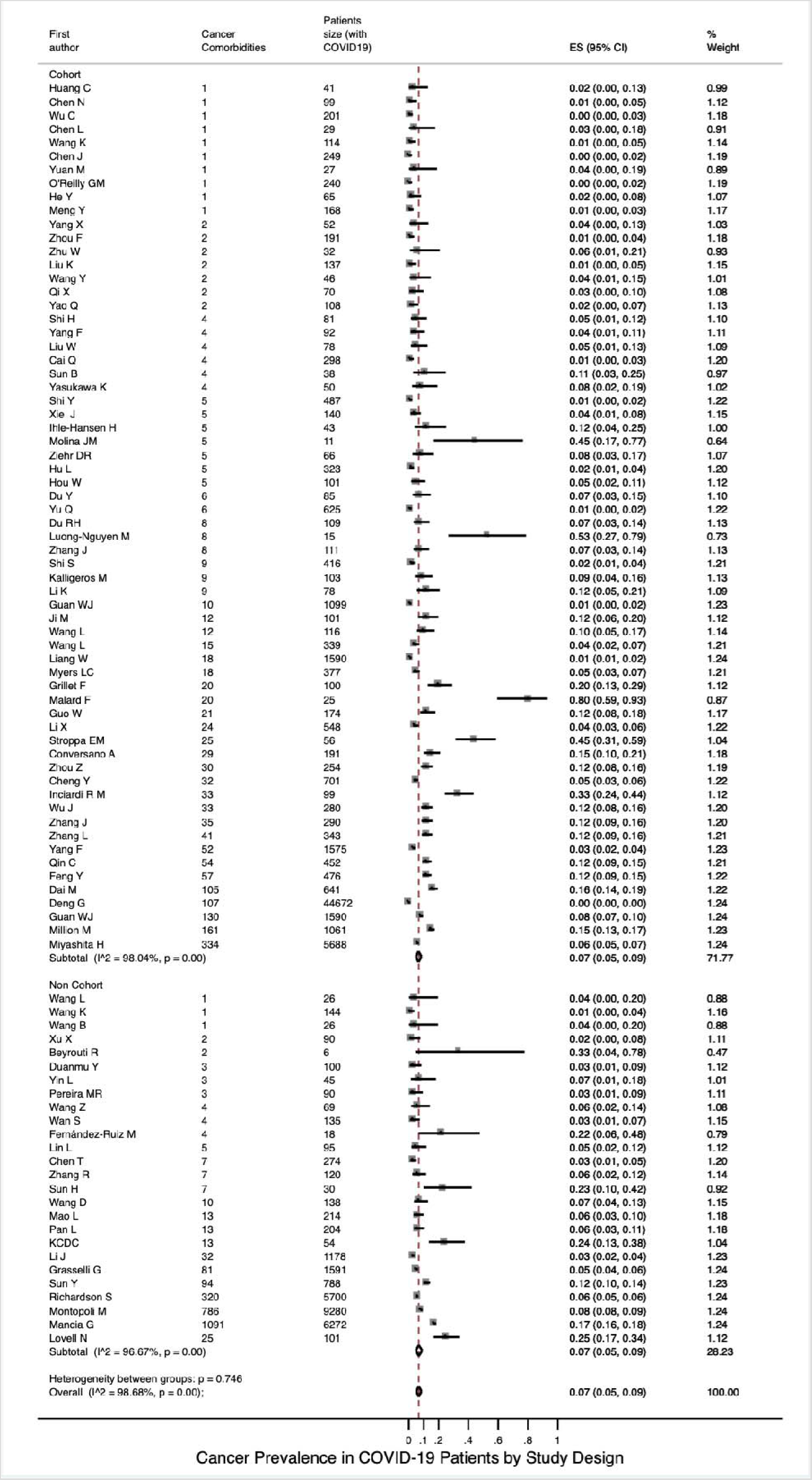
Cancer Prevalence in COVID-19 Patients by Study Design

### 3.3 Severe illness rate and mortality of COVID-19 Patients with Cancer Stratified by Continent

Based on the included 15 studies with available data, the pooled severe illness rate of COVID-19 patients with cancers was 0.33 (95% CI 0.26∽0.39; **Figure 8**). We also calculated the incidence of severe illness of COVID-19 patients without cancers based on 3 studies with available data (0.12, 95% CI 0.07∽0.17; **Figure S2 A**). Seventeen studies reported the death information for COVID-19 patients with cancers. Their mortalities were synthesized using a random-effects model and the pooled death rate was 0.18 (95% CI 0.14∽0.23; **Figure 9**). Based on the 5 included studies with available death data of COVID-19 patients without cancers, the pooled death rate was estimated to be 0.05 (95% CI 0.02∽0.08; **Figure S2 B**).

**Figure 8.**
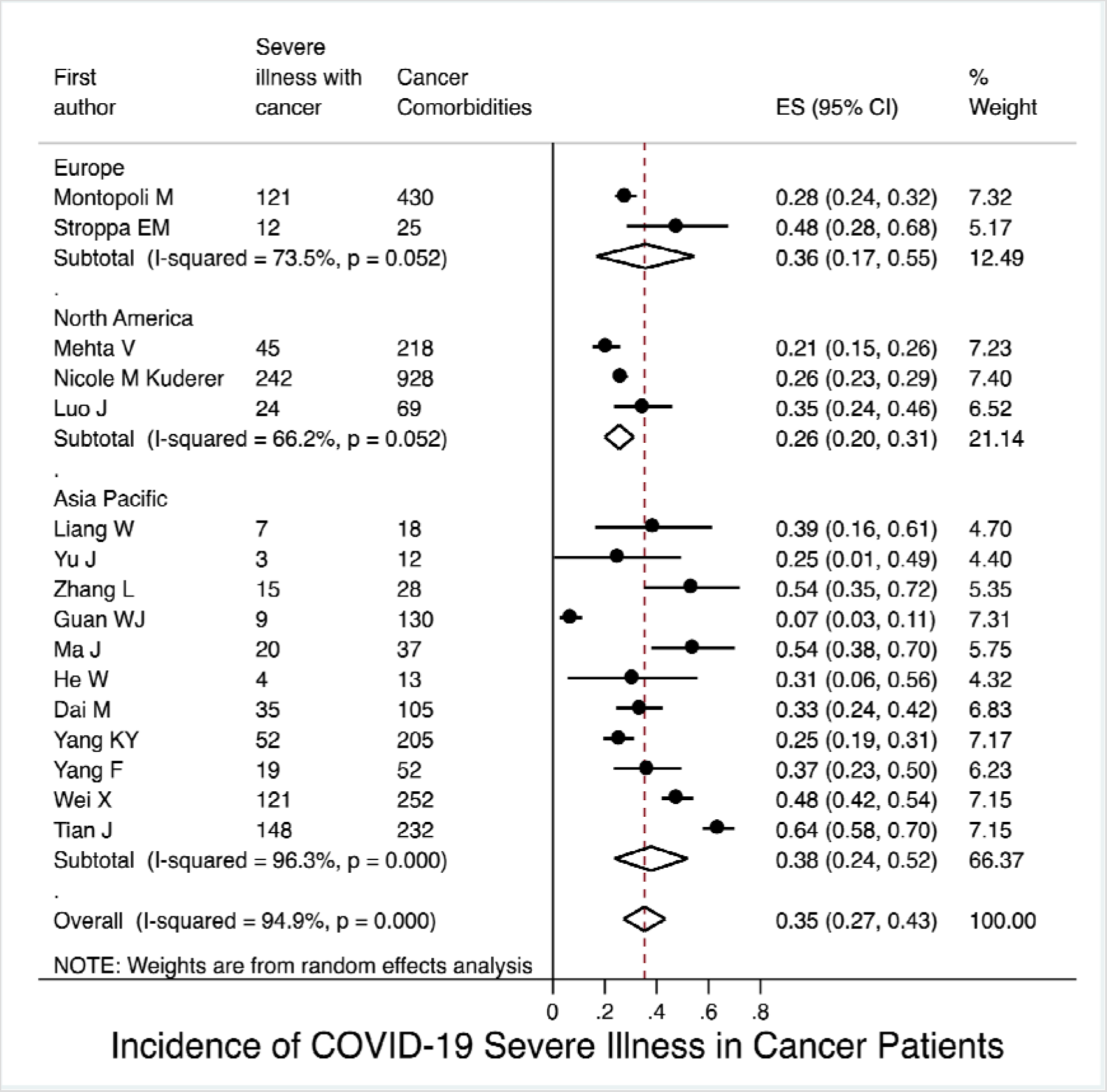
Incidence of Severe Illness among COVID-19 Patients with Cancer by Continents

**Figure 9.**
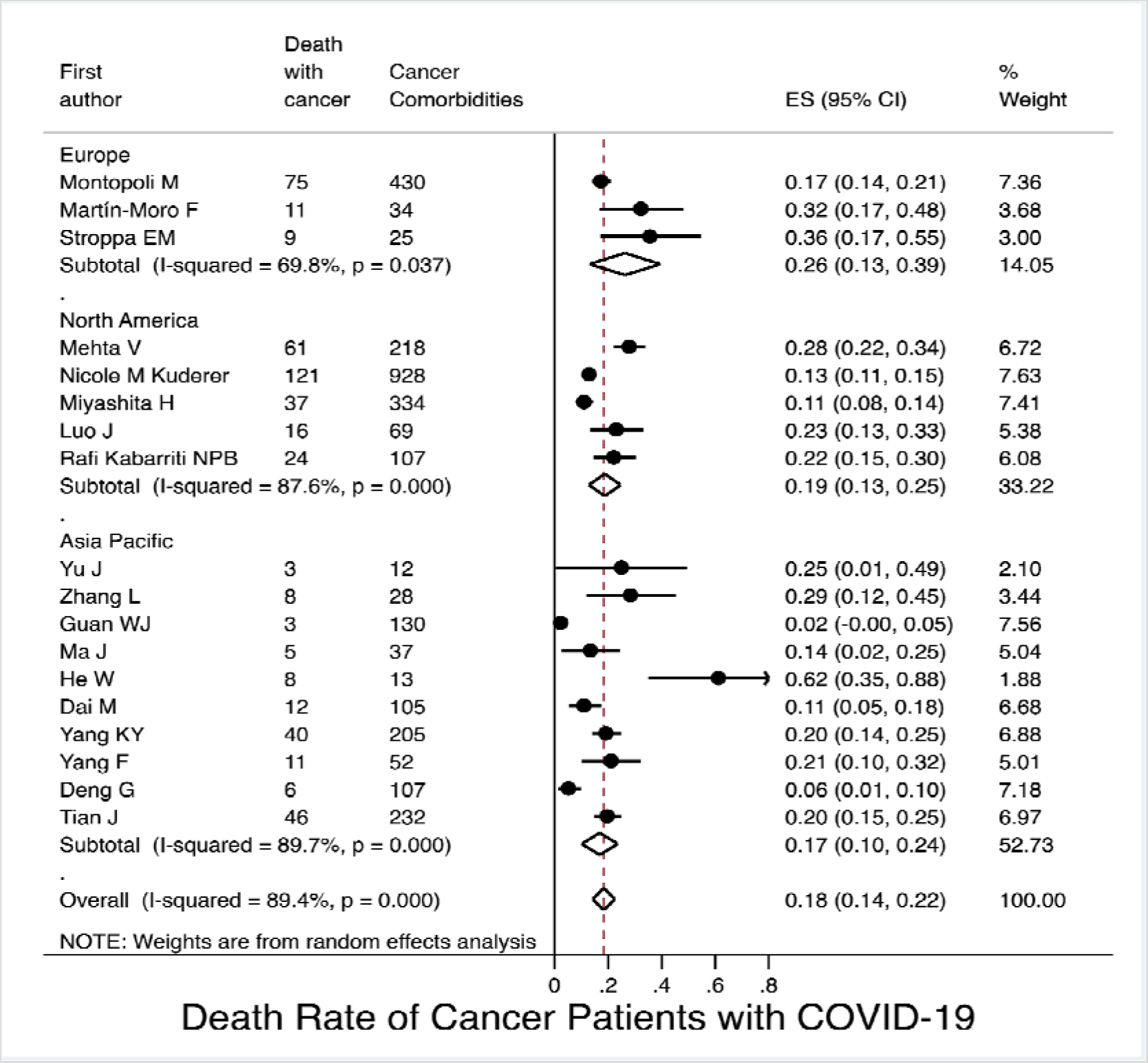
Death Rate of COVID-19 Patients with Cancer by Continents

For subgroup analysis of continents, from **Figure 8**, based on 10 studies with available data, the pooled incidence of severe illness of COVID-19 patients with cancers from Asia Pacific was 0.38 (95% CI 0.24∽0.52), similar to that from Europe based on 2 studies (0.36, 95% CI 0.17∽0.55); but was higher than that from North America based on 3 studies (0.26, 95% CI 0.20∽0.31). From **Figure 9**, the pooled death rate of COVID-19 patients with cancers with 10 studies was 0.17 for Asia Pacific (95% CI 0.10∽0.24), 0.26 for Europe with 3 studies (95% CI 0.13∽0.39), and 0.19 for North America with 5 studies (95%. CI 0.13∽0.25), respectively.

### 3.4 Heterogeneity, Meta-Regression, and Publication Bias Analysis

From the forest plots of the cancer prevalence among COVID-19 patients (**Figure 2∽7**), high heterogeneity was detected with the overall I^2^ = 98.68% (P< 0.001). **Table 3** showed the results of the metaregression for the included 90 studies for prevalence of cancer in COVID-19 patients. The between-study variance could be explained by the estimate difference by continents with statistically significant variable (I^2^=62.37%, τ^2^=0.0015, and P=0.04) as well as by countries (I^2^ = 46.74%, τ^2^ = 0.0011, and P<0.001). Age group (I^2^ = 70.49%), τ^2^ = 0.0016, and P = 0.20), study design (I^2^ = 57.22%, τ^2^ = 0.0015, and P = 0.35), and sample size (I^2^ = 70.17%, τ^2^ = 0.0015, and P = 0.075) may not explain the heterogeneity. There was no significant publication bias observed in this meta-analysis (funnel plot was shown in **Figure 10** with P = 0.224).

**Table 3.**
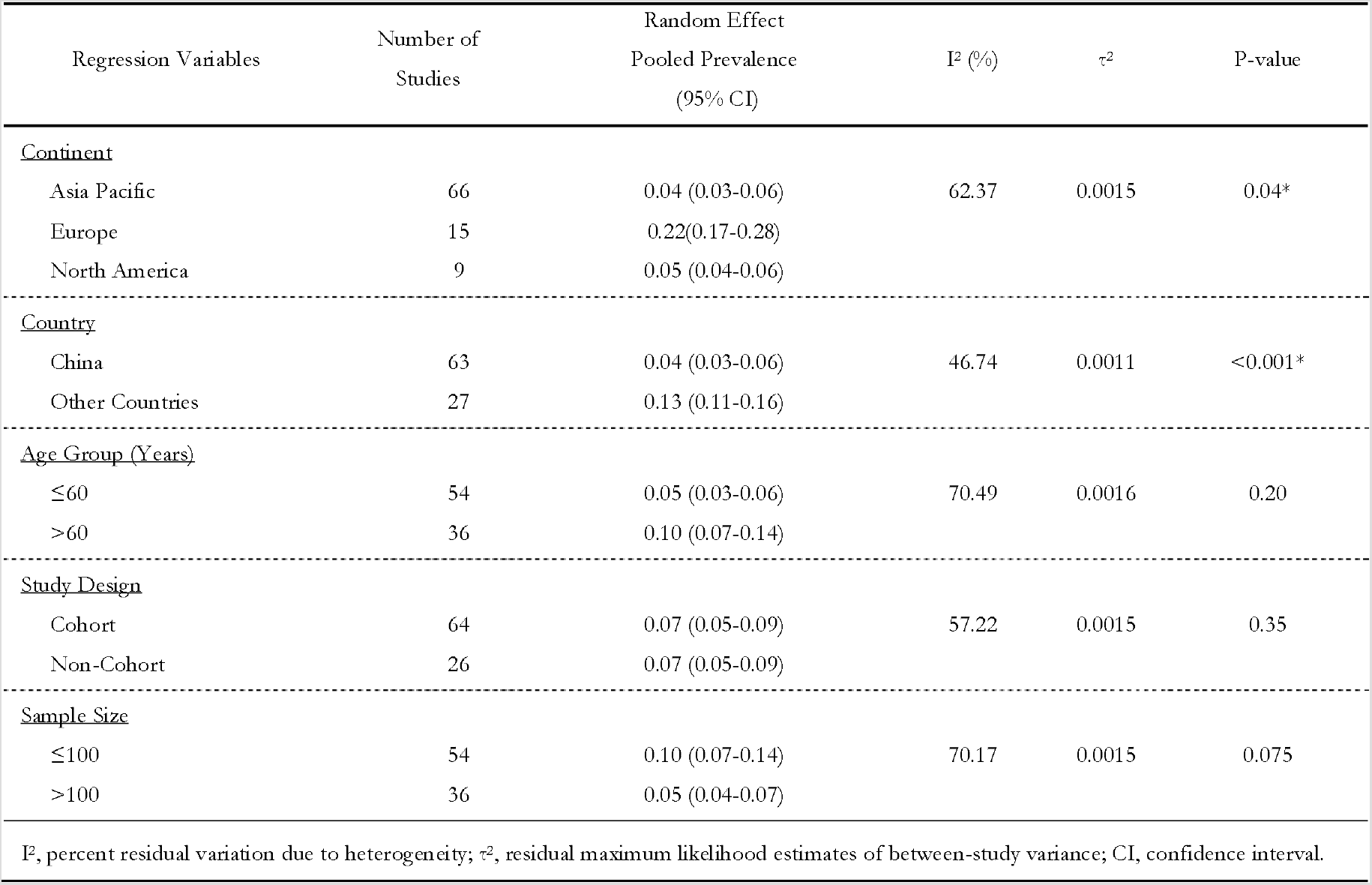
Summary of Meta Regression for the Included 90 studies.

| Table 3 Summary of Meta Regression for the Included 90 studies |  |  |  |  |  |
| --- | --- | --- | --- | --- | --- |
| Regression Variables | Number of Studies | Random Effect Pooled Prevalence (95% CI) | I<sup>2</sup> (%) | τ<sup>2</sup> | P-value |
| <u>Continent</u> |  |  |  |  |  |
| Asia Pacific | 66 | 0.04 (0.03-0.06) | 62.37 | 0.0015 | 0.04* |
| Europe | 15 | 0.22(0.17-0.28) |  |  |  |
| North America | 9 | 0.05 (0.04-0.06) |  |  |  |
| <u>Country</u> |  |  |  |  |  |
| China | 63 | 0.04 (0.03-0.06) | 46.74 | 0.0011 | <0.001* |
| Other Countries | 27 | 0.13 (0.11-0.16) |  |  |  |
| <u>Age Group (Years)</u> |  |  |  |  |  |
| ≤60 | 54 | 0.05 (0.03-0.06) | 70.49 | 0.0016 | 0.20 |
| >60 | 36 | 0.10 (0.07-0.14) |  |  |  |
| <u>Study Design</u> |  |  |  |  |  |
| Cohort | 64 | 0.07 (0.05-0.09) | 57.22 | 0.0015 | 0.35 |
| Non-Cohort | 26 | 0.07 (0.05-0.09) |  |  |  |
| <u>Sample Size</u> |  |  |  |  |  |
| ≤100 | 54 | 0.10 (0.07-0.14) | 70.17 | 0.0015 | 0.075 |
| >100 | 36 | 0.05 (0.04-0.07) |  |  |  |
| I<sup>2</sup>, percent residual variation due to heterogeneity; τ<sup>2</sup>, residual maximum likelihood estimates of between-study variance; CI, confidence interval. |  |  |  |  |  |

**Figure 10.**
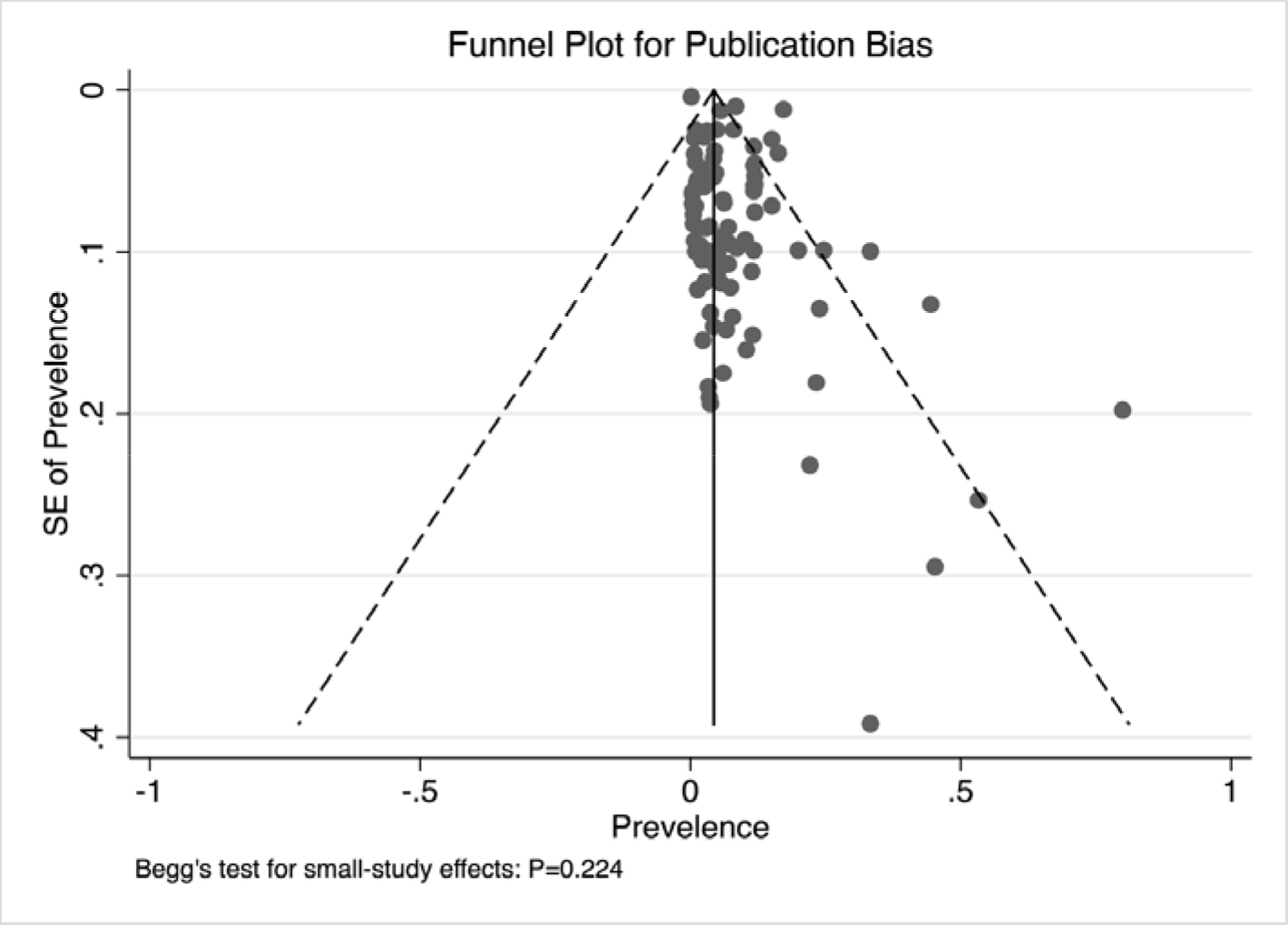
Funnel Plot with Pseudo 95% Confidence Limits and Begg’s Test

## 4 Discussion

This systematic review and meta-analysis of 107 global studies is the most comprehensive metaanalysis pooling formally published studies assessing the prevalence of COVID-19-infected cancer patients, and the incidence of severe illness and death rates in COVID-19-infected cancer patients. We showed that the overall pooled prevalence of cancer in patients with COVID-19 in these studies was 0.07, higher than Aakash et al.’s statistical result published in April 2020 (prevalence 0.02, 95% CI, 0.02∽0.03) [19]. In Asia-Pacific studies, prevalence was slightly lower at 0.04 than North America studies at 0.06, but in Europe studies, we found a significantly higher overall prevalence of 0.22. Moreover, studies from China showed a lower overall prevalence of 0.04 than other countries of 0.10. According to 2015 statistics [10], the incidence of cancer in the overall Chinese population (285.83 per 100 000 people [0·29%J), which is much lower than that of cancer in COVID-19-infected population in China.

In May 2020, a pre-printed meta-analysis by Venkatesulu et al., which included 31 studies and assessed outcomes in cancer patients affected by COVID-19 [20] demonstrated that cancer patients with COVID-19 had a higher likelihood of death (odds ratio, OR 2.54) and cancer patients were more likely to be intubated. They also showed that COVID-19-infected cancer patients were older than the normal population and had a higher proportion of comorbidities. However, this meta-analysis lacked pooled prevalence as a function of variables such as age and geographic region. We conducted a series of subgroup analyses of the prevalence from perspectives of continent, country, mean age, sample size, and study design type, providing us with more detailed information; and provided the pooled prevalence data of cancer morbidity among COVID-19 population. Interestingly, we found that age was associated with the prevalence of cancer in COVID-19 patients. Residence in Europe, Asia-Pacific or North America was also associated with differences in cancer prevalence. These geographical differences might be caused by the following reasons:

1. In response to the COVID-19 pandemic, many countries implemented policies like screening healthcare staff, and patients based on COVID-19 symptoms and travel history, placing restrictions on employee business travel, providing COVID-19 hotline and response teams, engaging in telehealth and online meetings, training staff in using personal protective equipment, and minimizing admissions and follow-up visits [19]. Both the prevalence and the clinical outcomes might be related to national or regional public health and epidemic prevention policies, which might also exert potential influence on the geographical differences. The situation in Asian countries including China, South Korea and Singapore may partly be due to the government’s policy, acting aggressively by using social-distancing measures to slow down disease spread, and performed extensive testing and isolating of infected people to stamp out potential transmission sources. This strategy helped the countries contain the outbreak. However, in comparison, several countries in Europe and North America took longer to deal with the spread of COVID-19. Some evidence suggests that those with multimorbidity including cancer were more susceptible to COVID-19 and more likely to be at risk of severe illness and poorer outcomes. Therefore, in cases where the disease has not been well controlled for a long time, the inadequate public health measures (such as not wearing a mask, premature large-scale gatherings) might cause cancer patients in these areas to be more likely to be infected with COVID-19.
2. The molecular subtypes of COVID-19 that are prevalent on different continents may have large differences in molecular structure, virulence, and invasiveness. In a phylogenetic network analysis of 160 COVID-19 genomes, Forster et al. found 3 central variants distinguished by amino acid alterations, which were named A, B, and C [21]. The A and C subtypes prevailed in significant proportions mainly in Europeans and Americans. By contrast, the B type was the most common type in East Asia, and its ancestral genome appears not to have spread outside East Asia without first mutating into derived B types, pointing to founder-effects or immunological or environmental resistance against this type outside Asia [21].
3. There are inequalities in COVID-19 susceptibility and clinical outcomes between people from different ethnic backgrounds. There has been debate about the extent to which COVID-19 affects ethnic groups differently and related studies so far have explored the impact of ethnicity on COVID-19 mortality and morbidity, for instance, Santorelli et al. analyzed the mortality rates in 1,276 inpatients in Bradford with test results for COVID-19 by ethnic group [22]. The age-adjusted risk of dying from COVID-19 was slightly lower in South Asian compared to British patients (RR 0.87; 95% CI 0.41∽1.84) [22]. Similarly, Public Health England (PHE) reported on the disparities in the risk and outcomes of COVID-19 [23]. After adjusting for sex, age, deprivation, and region, people from a Black, Asian, and Minority Ethnic (BAME) background had a higher risk of death from COVID-19 than British people, although once comorbidities were accounted for, there was no difference in COVID-19 mortality between ethnic groups [23]. Few studies have compared the prevalence or incidence rate of COVID-19 between ethnic groups. In May 2020, Niedzwiedz et al. linked participants in the UK Biobank [24] to COVID-19 testing data from PHE. Of 392,116 participants in the cohort, 2,658 were tested for COVID-19, of whom 948 tested positive. The positive incidences were higher in Irish (RR 1.42; 95% CI 1.00∽2.03), South Asian (RR 2.42; 95% CI 1.75∽3.36) and Black (RR 3.35; 95% CI 2.48∽4.53) population than the British population [24]. Our meta-analysis indicated that the Caucasian patients with cancer seemed to be more likely to be infected with COVID-19 than the Asian population in contrast to existing epidemiological studies.

In May 2020, Kuderer et al. published a cohort study based on 928 COVID-19-infected cancer patients [8]. They found that independent factors associated with mortality include age, male sex, smoking status, number of comorbidities, Eastern Cooperative Oncology Group performance status of 2 or higher, active cancer, and receipt of azithromycin plus hydroxychloroquine. In our study, because the data of potential risk factors associated with mortality was limited among the included studies, we did not assess the association between the clinical outcome and potential prognostic variables mentioned above, a limitation of this study. However, one of the novelties and strengths of the present meta-analysis is that besides the prevalence, we also performed the subgroup analysis as a function of continent for incidence of severe illness and death rate among COVID-19 patients with cancers. There were some interesting findings (**Figure 11**): *1)* the European COVID-19 patients had both the highest cancer prevalence (0.22) and cancer patient mortality (0.26); *2)* the North American COVID-19 patients had a similar cancer prevalence as Asia Pacific patients, but had the lowest cancer patient severe illness rate (0.26); *3)* compared with Asia Pacific, the European COVID-19 patients had a much higher cancer prevalence, but their cancer patient severe illness rates were similar (Asia Pacific 0.35 vs. Europe 0.36); and *4)* compared with Asia Pacific, the North American COVID-19 patients had a similar cancer prevalence and cancer patient mortality, but the incidence of severe illness of cancer patients (0.26) was much lower. Overall, the European COVID-19 patients seemed the most likely to both develop cancer and progress to severe illness and death (for COVID-19 patients with cancers). Although the Asia Pacific COVID-19 patients had the lowest cancer prevalence, their severe illness rate was similar as European’s, Finally, we also performed a subgroup analysis of severe illness rate and mortality by continent among COVID-19 patients without cancers (data were shown in **Figure S2 A-B**), giving that the included studies were limited, we did not perform subgroup analysis.

**Figure 11.**
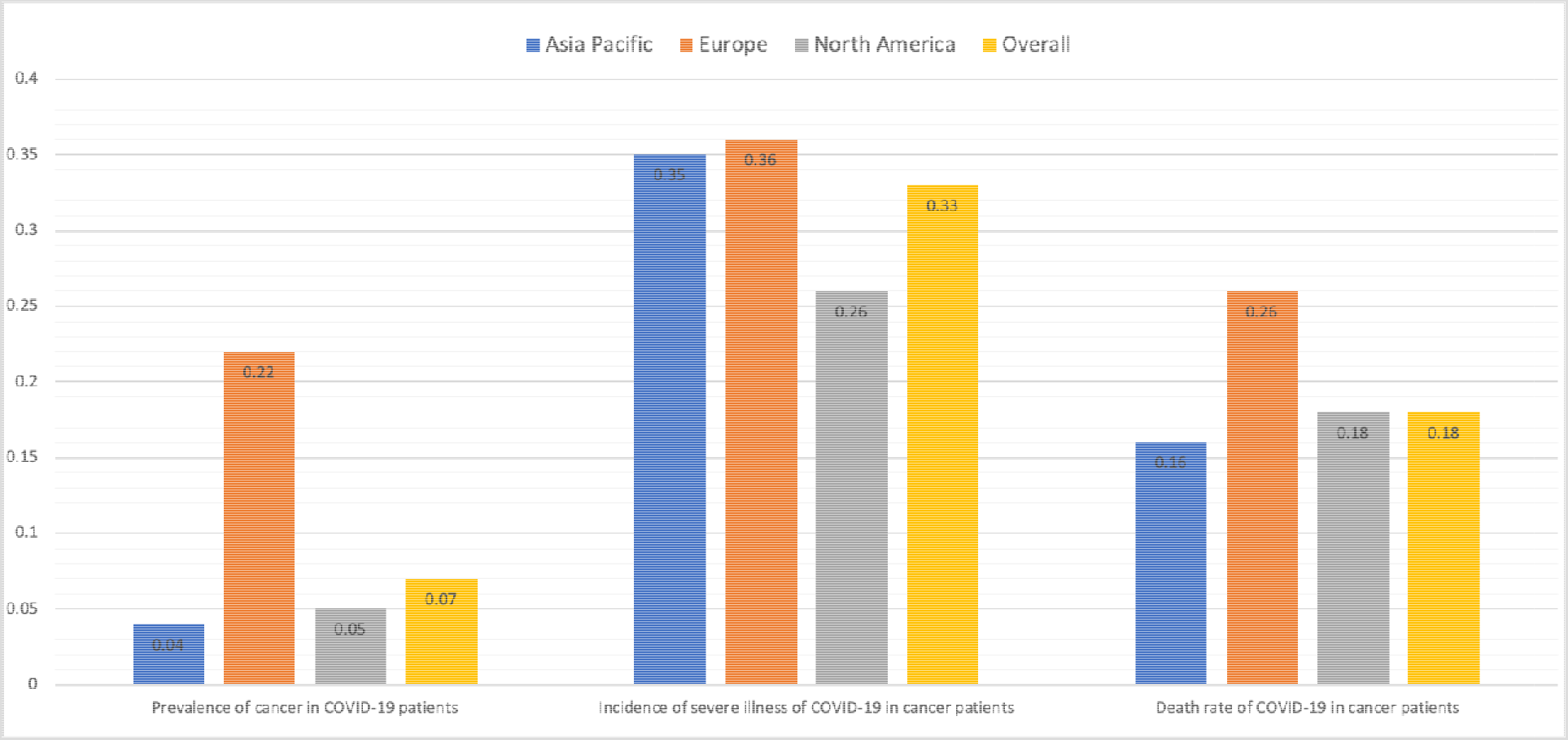
Prevalence, Severe Illness Rate and Death Rate of COVID-19 Patients with Cancer by Continents

Another strength of this meta-analysis is that we descriptively demonstrate the incidence of severe illness and death rate in COVID-19 patients with cancers and without cancers. Cancer patients usually have systemic immunosuppressive states caused by the malignancy itself and anticancer treatments, such as chemotherapy, surgery, or immunomodulatory drugs like PD-1/PD-L1 inhibitors [4, 6, 9], thus their ability to resist virus virulence and invasiveness is relatively weaker. In addition, they are often at older ages with one or more major comorbidities, putting them at increased risk for COVID-19-related mortality [7]. Our systematic review and meta analysis had several limitations: *1)* Heterogeneity was observed in the included studies both for the prevalence estimation and for the severe illness and mortality analysis. We minimized heterogeneity influence by using the random effects model and also performed exploratory subgroup analyses based on age, continent, country, sample size, and study design type, as well as we performed both metan and metaprop to test the robustness of the results. *2)* This study might be potentially limited by the retrospective nature of most of the included studies. To minimize possible inaccuracies, we conducted subgroup analysis by study design type, respectively pooling the prevalence in cohort studies and non-cohort studies (case-series studies, case-control studies, and cross-sectional studies). *3)* The definitions of “severe illness” were not unitary and described differently across the included studies, and this might lead to significant heterogeneity. Generally, severe illness referred to a composite of severe illness requiring mechanical ventilation, admission to an ICU, admission to hospital, or a combination of these; mechanical ventilation; admission to an ICU; admission to hospital; and need for supplemental oxygen during the course of COVID-19. *4)* Most included studies did not give a clearly specified time period for the deathate; hence the objectivity of the mortality comparison might be influenced by possible reporting bias due to the time unconformity. In addition, longer-term follow up and larger sample sizes were needed to more completely understand the epidemiological and clinical characteristics of cancer patients in COVID-19 infection. *5)* When performing the subgroup analysis for severe illness rate and mortality by continent, the included studies for each subgroup were limited due to data availability.

## 5 Conclusions

Taken together with previously published results, our meta-analysis provides a comprehensive picture of epidemiological and clinical characteristics of cancer patients in COVID-19 infection. The overall cancer prevalence among COVID-19 patients was estimated as 0.07 (95% CI 0.05∼0.09) The prevalence increased with age. The prevalence in Europe was much higher than that in Asia Pacific and North America. The COVID-19 patients with cancer were at-risk for a more severe illness and a higher death rate. This study of COVID-19-infected cancer patients reinforced important considerations for clinical care and emphasized the urgent needs for more data with longer term follow-up, larger sample sizes, and more detailed sociodemographic and clinicopathological variables. In the future, as more data becomes available, it will like to investigate differences by sociodemographic and clinicopathological features (like age, sex, race, smoking status, symptom and sign, cancer type, laboratory results, and tumor stage, etc.) between COVID-19-infected patients with cancer and without cancer.

## Data Availability

The data were abstracted from the published paper.

## Supplemental Files

**Table S1.** The Search Strategy and Search Results

| **A Medline Search** |  |  |
| --- | --- | --- |
| Entry | Medline Search Strategy | Results |
| 1 | (cancer or cancers or tumor or tumors or neoplasm or neoplasms or adenoma or adenomas or carcinoma or carcinomas).mp. [mp=title, abstract, original title, name of substance word, subject heading word, floating sub-heading word, keyword heading word, organism supplementary concept word, protocol supplementary concept word, rare disease supplementary concept word, unique identifier, synonyms] | 3920545 |
| 2 | (2019-ncov or novel coronavirus or COVID-19 or SARS-CoV-2).mp. [mp=title, abstract, original title, name of substance word, subject heading word, floating sub-heading word, keyword heading word, organism supplementary concept word, protocol supplementary concept word, rare disease supplementary concept word, unique identifier, synonyms] | 7450 |
| 3 | #1 AND #2 | 258 |
| **B Embase Search** |  |  |
| Entry | Embase Search Strategy | Results |
| 1 | (cancer or cancers or tumor or tumors or neoplasm or neoplasms or adenoma or adenomas or carcinoma or carcinomas).mp. [mp=title, abstract, original title, name of substance word, subject heading word, floating sub-heading word, keyword heading word, organism supplementary concept word, protocol supplementary concept word, rare disease supplementary concept word, unique identifier, synonyms] | 5463874 |
| 2 | (2019-ncov or novel coronavirus or Covid-19 or SARS-CoV-2).mp. [mp=title, abstract, original title, name of substance word, subject heading word, floating sub-heading word, keyword heading word, organism supplementary concept word, protocol supplementary concept word, rare disease supplementary concept word, unique identifier, synonyms] | 6534 |
| 3 | #1 AND #2 | 285 |
| **C Web of Science Search** |  |  |
| Entry | Web of Science Search Strategy | Results |
| 1 | TS= (cancer or cancers or tumor or tumors or neoplasm or neoplasms or adenoma or adenomas or carcinoma or carcinomas) | 3,731,583 |
| 2 | TS=(2019-ncov or novel coronavirus or covid-19 or SARS-CoV-2) | 3,289 |
| 3 | #1 AND #2 | 94 |
| **D Cochrane Library Search** |  |  |
| Entry | Cochrane Library Search Strategy | Results |
| 1 | cancer or cancers or tumor or tumors or neoplasm or neoplasms or adenoma or adenomas or carcinoma or carcinomas | 211213 |
| 2 | “novel coronavirus” or “COVID-19” | 68 |
| 3 | #1 AND #2 | 8 |
| 4 | Cochrane Central Register of Controlled Trials | 3 |

**Figure S1(A).**
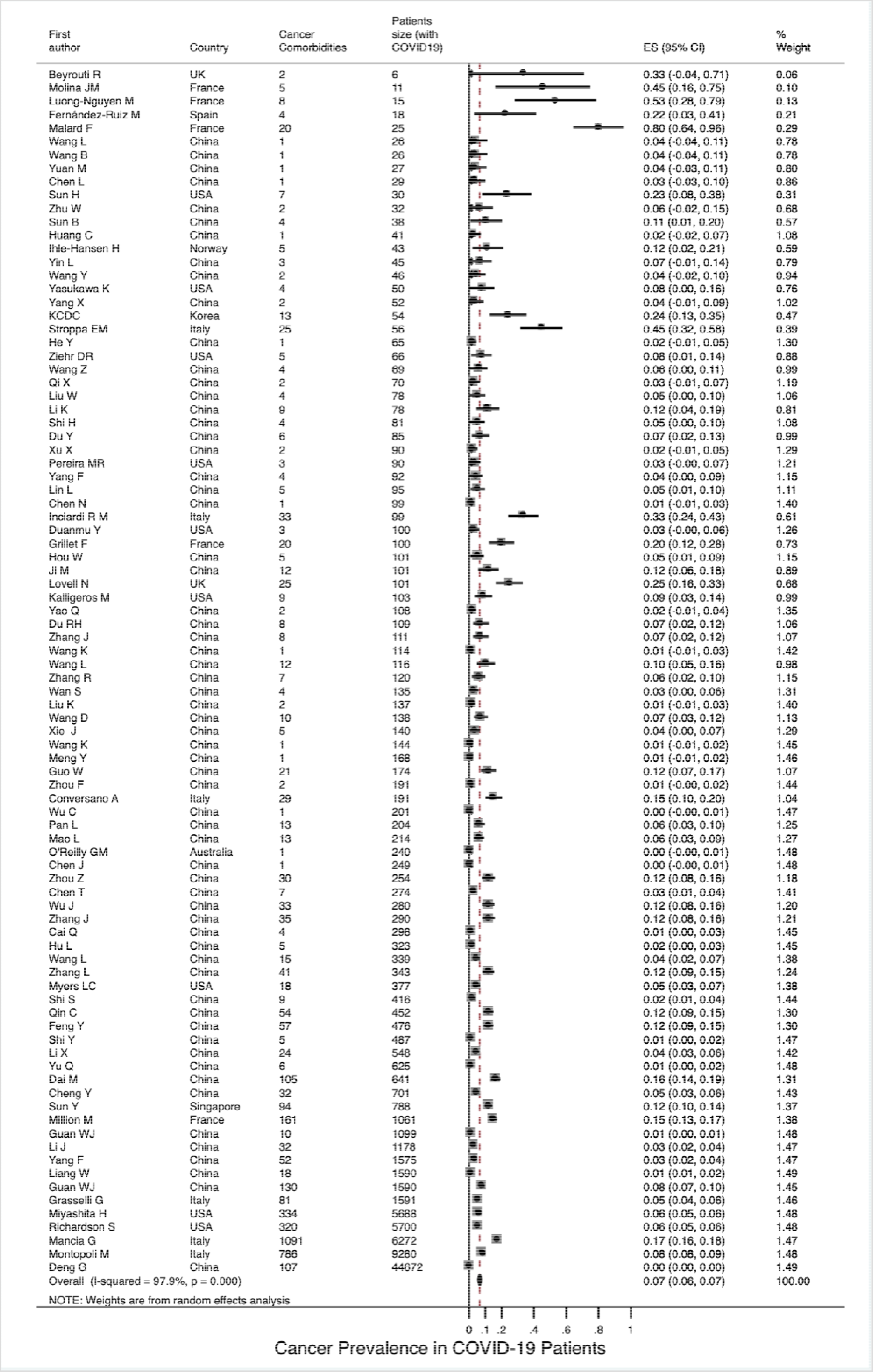
Overall Cancer Prevalence in COVID-19 Patients Analyzed by Metan Method

**Figure S1(B).**
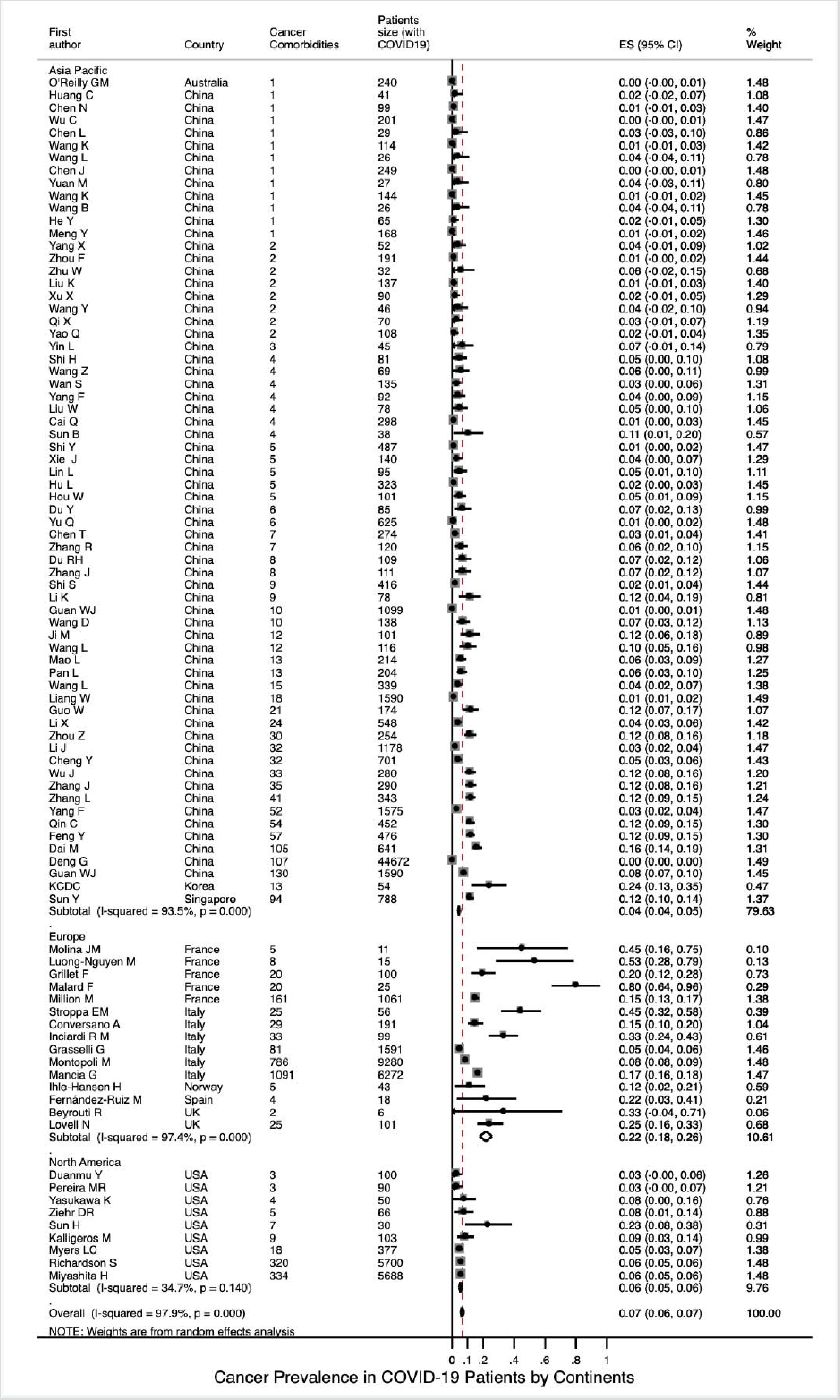
Cancer Prevalence in COVID-19 Patients by Continents Analyzed by Metan Method

**Figure S1(C).**
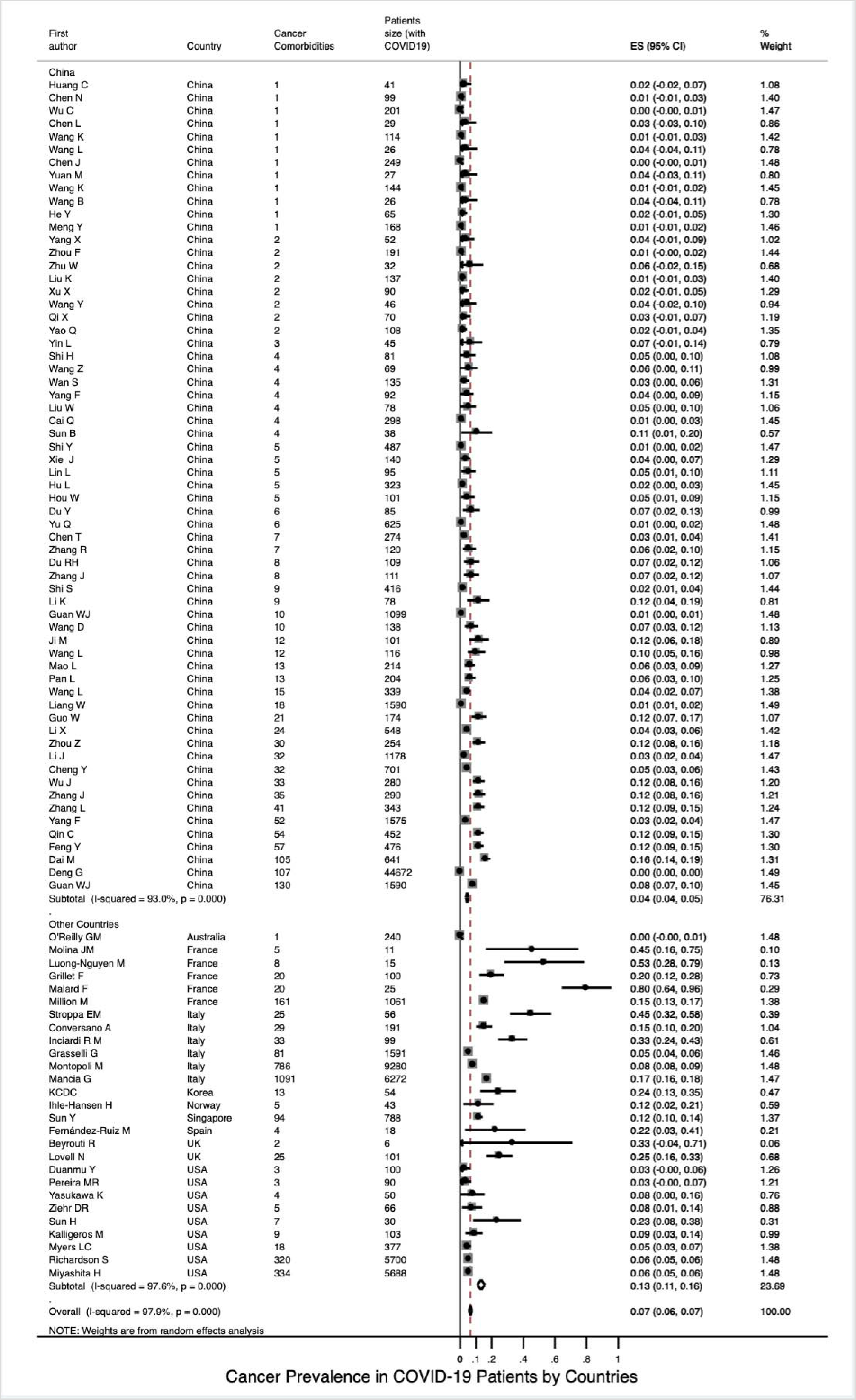
Cancer Prevalence in COVID-19 Patients by Countries Analyzed by Metan Method

**Figure S1(D).**
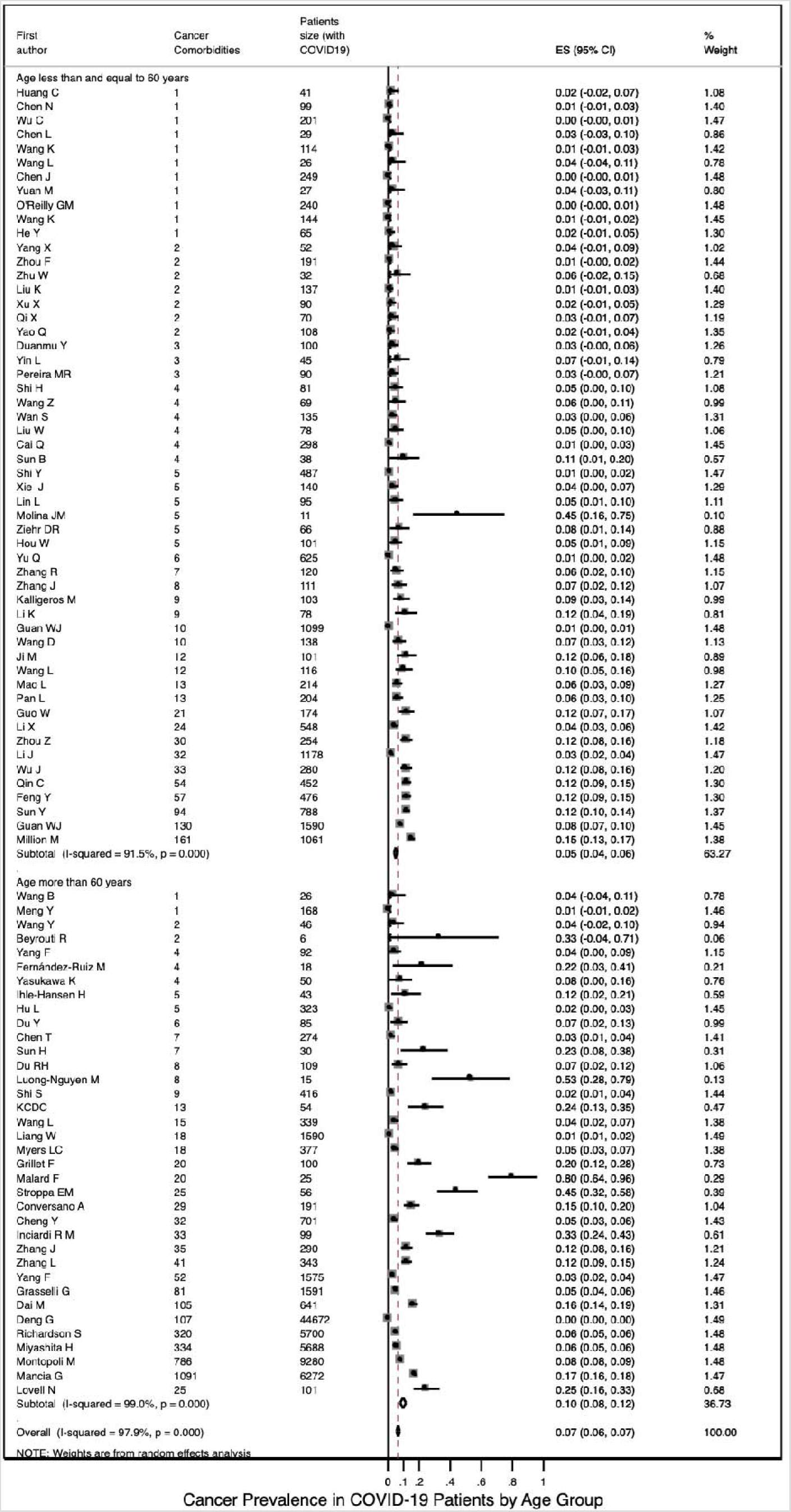
Cancer Prevalence in COVID-19 Patients by Age Group Analyzed by Metan Method

**Figure S1(E).**
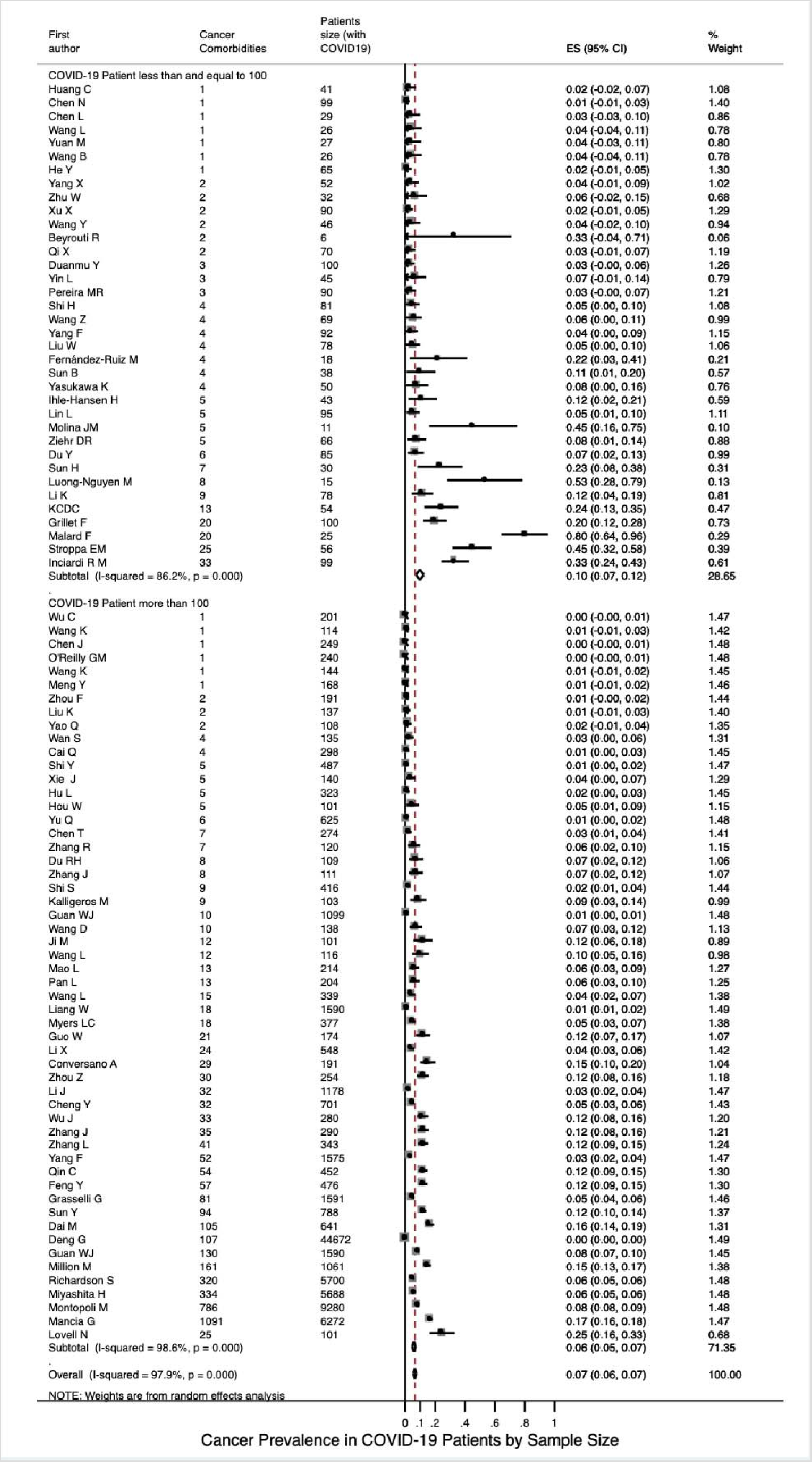
Cancer Prevalence in COVID-19 Patients by Sample Size Analyzed by Metan Method

**Figure S1(F).**
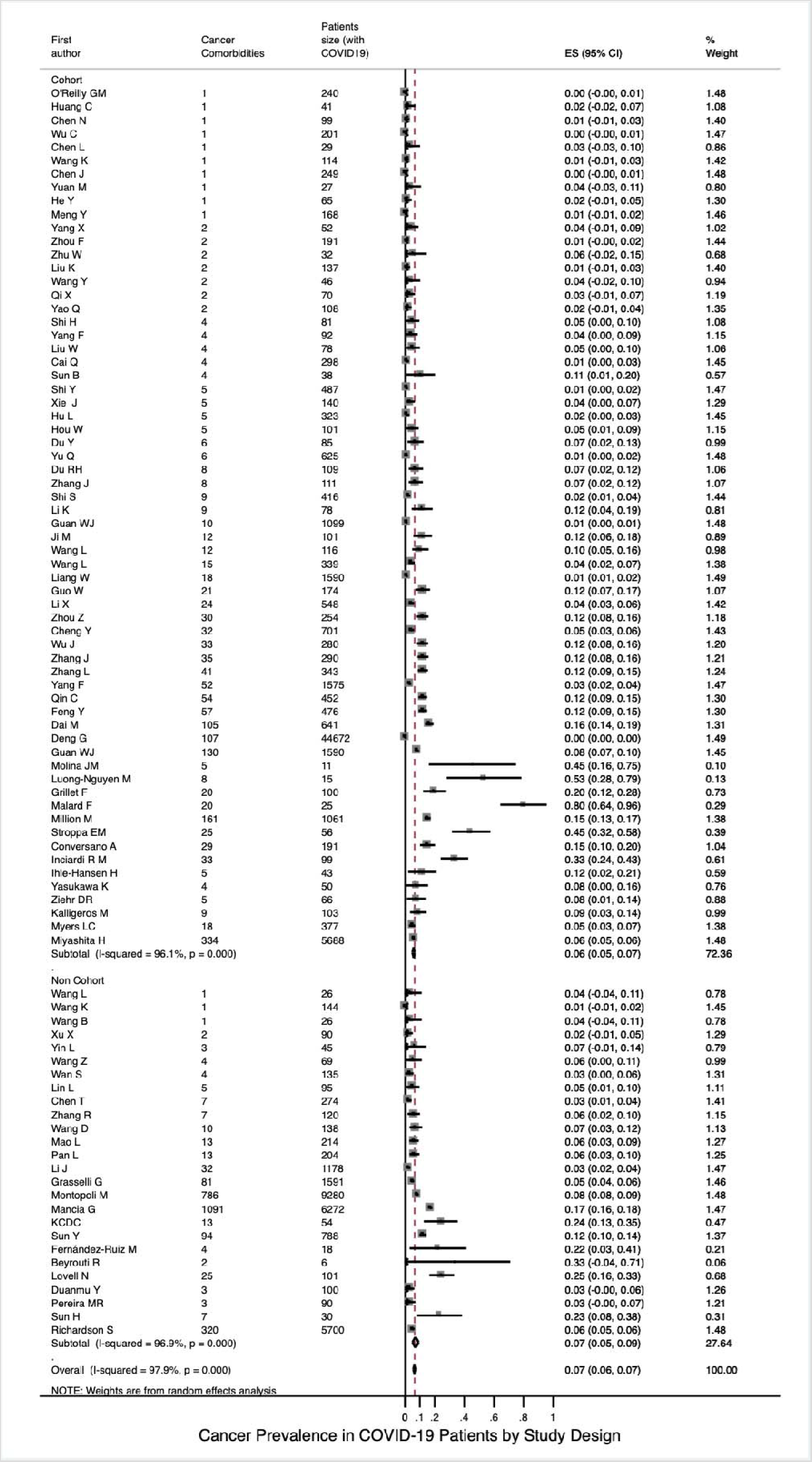
Cancer Prevalence in COVID-19 Patients by Study Design Analyzed by Metan Method

**Figure 2(A).**
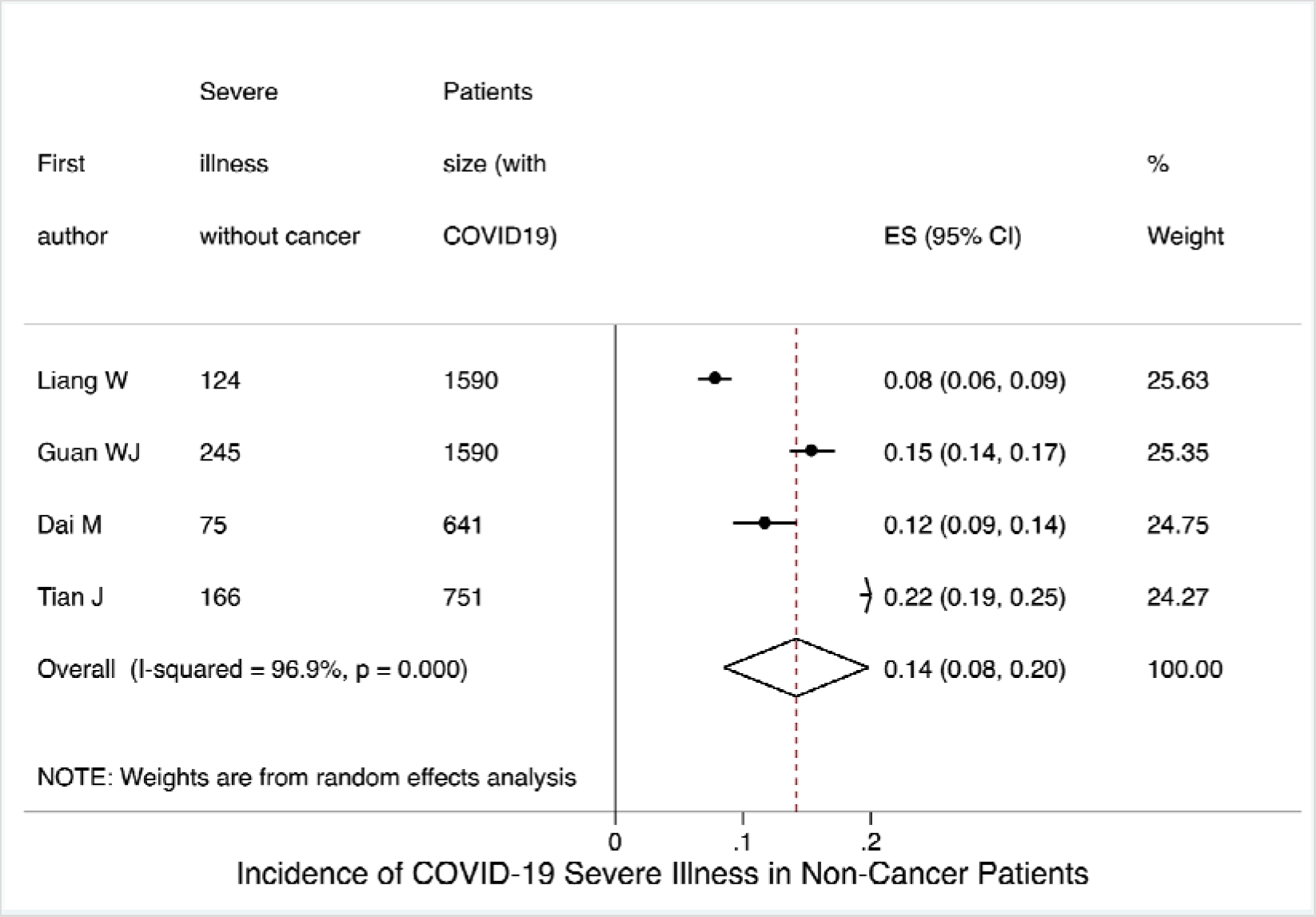
Incidence of Severe Illness with COVID-19 in Non-Cancer Patients

**Figure 2(B).**
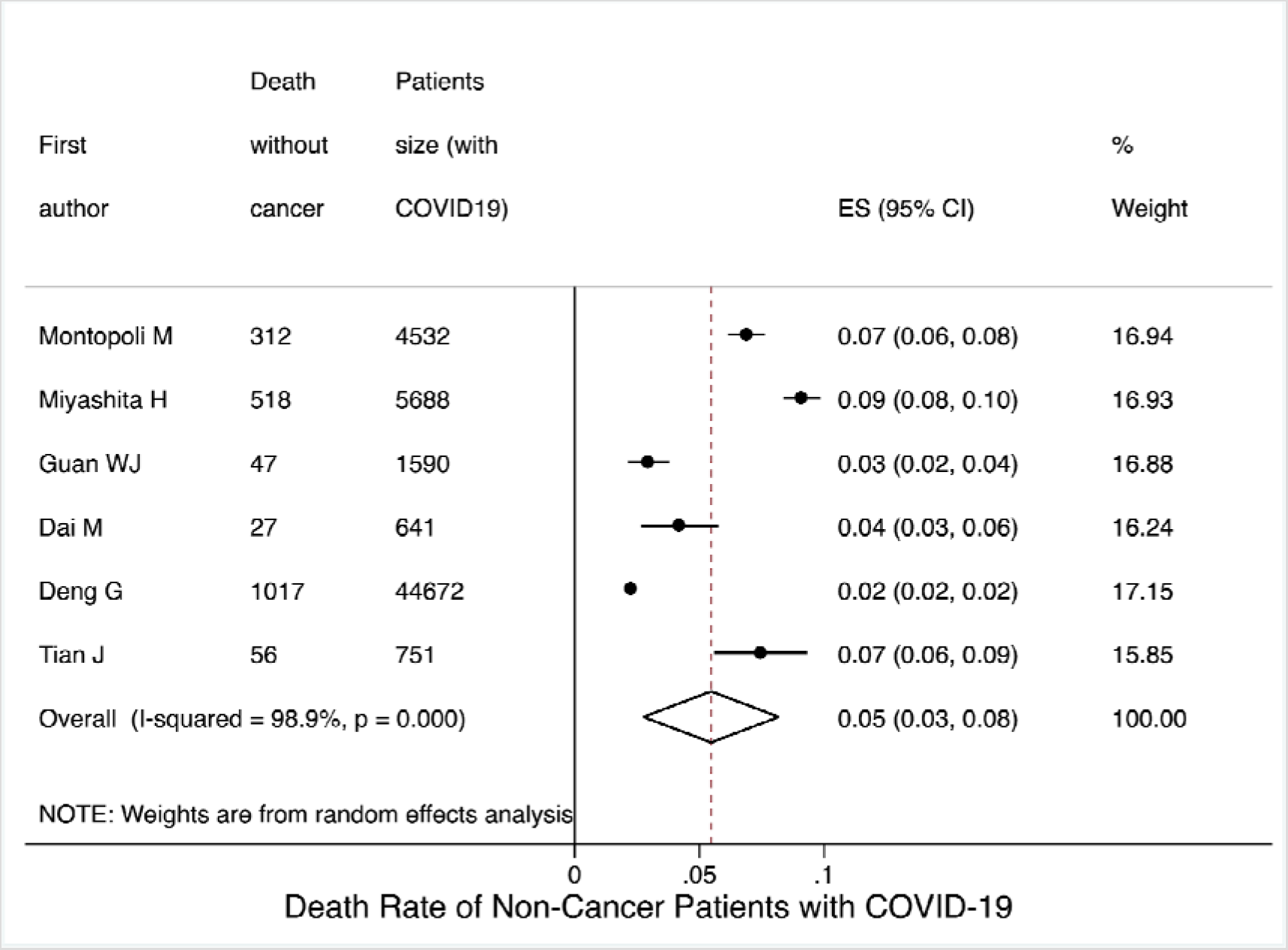
Death Rate of Non-Cancer Patients with COVID-19

## Acknowledgments

None.

## Author Contributions Conceptualization

Yu Jiang, Lin Zhang, Xiangyi Kong, Yi Fang, Jing Wang; Data Collection, Yihang Qi, Lin Zhang, Xiangyi Kong; Methodology, Junjie Huang, Yang Zhao, Xuzhen Qin, Zhihong Qi, Adejare (jay) Atanda, Lei Zhang, Peng Jia; Supervision, Yu Jiang, Asieh Golozar; Writing – original draft, Xiangyi Kong, Lin Zhang, Yihang Qi; Writing – review & editing, Asieh Golozar, Adejare (jay) Atanda, Lei Zhang, Xiangyi Kong, Lin Zhang. All authors have read and agreed to the published version of the manuscript.

## Ethical Approval

The analysis followed the Preferred Reporting Items for Systematic Reviews and Meta-Analyses (PRISMA) guidelines. National Cancer Center/National Clinical Research Center for Cancer/Cancer Hospital, Chinese Academy of Medical Sciences and Peking Union Medical College institutional review board approved this study as exempt.

## Registration

The research protocol has been registered and approved in PROSPERO. The registration number is CRD42020196014.

